# Monitoring SARS-CoV-2 in wastewater during New York City’s second wave of COVID-19: Sewershed-level trends and relationships to publicly available clinical testing data

**DOI:** 10.1101/2022.02.08.22270666

**Authors:** Catherine Hoar, Francoise Chauvin, Alexander Clare, Hope McGibbon, Esmeraldo Castro, Samantha Patinella, Dimitrios Katehis, John J. Dennehy, Monica Trujillo, Davida S. Smyth, Andrea I. Silverman

## Abstract

New York City’s ongoing wastewater monitoring program tracked trends in sewershed-level SARS-CoV-2 loads starting in the fall of 2020, just before the start of the City’s second wave of the COVID-19 outbreak. During a five-month study period, from November 8, 2020 to April 11, 2021, viral loads in influent wastewater from each of New York City’s 14 wastewater treatment plants were measured and compared to new laboratory-confirmed COVID-19 cases for the populations in each corresponding sewershed, estimated from publicly available clinical testing data. We found significant positive correlations between viral loads in wastewater and new COVID-19 cases. The strength of the correlations varied depending on the sewershed, with Spearman’s rank correlation coefficients ranging between 0.38 and 0.81 (mean = 0.55). Based on a linear regression analysis of a combined data set for New York City, we found that a 1 log_10_ change in the SARS-CoV-2 viral load in wastewater corresponded to a 0.6 log_10_ change in the number of new laboratory-confirmed COVID-19 cases/day in a sewershed. An estimated minimum detectable case rate between 2 - 8 cases/day/100,000 people was associated with the method limit of detection in wastewater. This work offers a preliminary assessment of the relationship between wastewater monitoring data and clinical testing data in New York City. While routine monitoring and method optimization continue, information on the development of New York City’s ongoing wastewater monitoring program may provide insights for similar wastewater-based epidemiology efforts in the future.

## Introduction

In March 2020, New York City became an epicenter of the coronavirus disease 2019 (COVID-19) pandemic. In response to this first wave of COVID-19 cases, the New York City Department of Environmental Protection (NYC DEP) - the city agency responsible for wastewater collection and treatment - launched a wastewater monitoring program with the goal of tracking sewershed-level trends in the concentration of Severe Acute Respiratory Syndrome Coronavirus 2 (SARS-CoV-2), the virus that causes COVID-19. The program was developed in partnership with researchers at New York University, Queens College, Queensborough Community College, and The New School, with all routine analysis conducted in the NYC DEP’s existing microbiology laboratory under the management of the NYC DEP.

Wastewater-based epidemiology (WBE) programs for COVID-19, including the one in New York City (NYC), were established on the premise that SARS-CoV-2 virions are excreted in the human waste of individuals infected with SARS-CoV-2 and that the resulting concentrations of viral RNA measured in wastewater are indicative of disease incidence or prevalence in the contributing sewershed. Significant associations between SARS-CoV-2 RNA concentrations measured in wastewater and metrics of COVID-19 disease incidence--including case rates--have been shown at scales ranging from single buildings to entire sewersheds.^1–3^ Early reports from WBE programs suggested promising predictive applications that could help inform COVID-19 response measures,^4,5^ sparking widespread interest in SARS-CoV-2 monitoring programs around the world.^6,7^ While the extent to which wastewater data is a leading indicator of trends in COVID-19 incidence ahead of clinical data may vary depending on clinical testing rates,^8,9^ WBE data do offer the advantage of providing information representative of entire populations, free from clinical testing-related biases. In NYC, where communities of color and high-poverty areas were disproportionately impacted by the first wave of the COVID-19 pandemic,^10^ testing rates varied spatially, with significant demographic-based disparities.^11^ In situations where clinical testing does not adequately sample vulnerable populations, WBE may help inform modifications to testing strategies and provide supplemental information regarding COVID-19 trends. Wastewater monitoring is therefore a potential tool to identify new outbreaks of COVID-19 after high clinical testing rates associated with major “waves” of disease incidence have subsided or when resources and technical capacity for extensive clinical testing of individuals are limited.

These opportunities make WBE an attractive option for many municipalities, including NYC, to confirm findings from clinical testing about population-level COVID-19 dynamics and to monitor for new outbreaks in instances when testing is inadequate. In August 2020, the NYC DEP’s SARS-CoV-2 wastewater monitoring program began routine analysis of influent wastewater collected from NYC’s 14 wastewater treatment plants (referred to as wastewater resource recovery facilities (WRRF) by the NYC DEP) (SI Table S1), capturing data during the region’s second wave of COVID-19 cases, which started in the fall of 2020. The sewershed catchment areas contributing to each of the 14 WRRFs vary markedly in size, serving populations ranging from approximately 120,000 to 1.2 million residents. To assess the relationship between NYC sewershed-level SARS-CoV-2 RNA concentrations and confirmed cases of COVID-19 within each sewershed, wastewater data were compared to publicly available case data provided by the NYC Department of Health and Mental Hygiene (DOHMH). In presenting findings from the NYC DEP, we also aim to provide insights into the development of a sustainable wastewater monitoring program designed for long-term, routine tracking of trends in virus loads for multiple sewersheds serving a large urban population.

## Methods

### Sample collection and processing

24-h flow-weighted composite influent wastewater samples were collected from each of NYC’s 14 WRRFs twice weekly beginning August 31, 2020. From January 31, 2021 to April 18, 2021 sampling was reduced to once weekly. Each composite sample consisted of eight grab samples collected every three hours beginning at 7:00 AM on the sampling date. Samples were transported on ice and stored at 4 °C until processing, which started within twelve hours after the final grab sample was collected. For each sampling date, one of the 14 samples was analyzed in duplicate and the remainder were analyzed as single samples; facilities were selected for duplicate analysis on a rotating basis. A method blank containing Type I deionized water was included with each set of samples to confirm the absence of contamination during sample processing. Detailed descriptions of materials, methods, and data analysis are provided in the SI. In brief, 40-mL aliquots of the 24-h composite samples were first pasteurized (60 ºC, 90 min), and then centrifuged (5000 x *g*, 4 ºC, 10 min) to remove solids. The supernatant was filtered (0.22 µm, cellulose acetate) and then subjected to virus concentration using polyethylene glycol (PEG) precipitation (addition of 4.0 g PEG and 0.9 g NaCl followed by overnight incubation at 4 ºC, and centrifugation at 12,000 x *g* at 4 ºC for 120 min to pellet viruses).^12^ The supernatant was discarded and RNA was extracted from the concentrated PEG pellet using the Qiagen QiaAmp Viral RNA Mini Kit with modifications (described in the SI).

### SARS-CoV-2 quantification by RT-qPCR

A one-step RT-qPCR assay was used to quantify copies of the SARS-CoV-2 nucleocapsid (N) gene, targeting the N1 region (CDC RUO Primers and Probes, Integrated DNA Technologies^13^) in triplicate reactions on a StepOnePlus Real-Time PCR System (Thermo Fisher Scientific). Synthetic SARS-CoV-2 RNA covering > 99.9% of the viral genome (Twist Bioscience Control 1, GENBANK ID MT007544.1) served as both a positive control and standard used in a decimal serial dilution for quantification of N1 gene copies.

The limit of detection (LOD) and limit of quantification (LOQ) for the assay were estimated from replicate standard curves as described by Forootan et al. 2017^14^ and found to be 4,500 copies/L and 15,000 copies/L, respectively. Note that these LOD and LOQ values as well as calculated sample concentrations are relative to the approximate concentration of the synthetic RNA control reported by the manufacturer, as absolute quantification of the RNA control was not feasible when sample analysis began. Note that quantification of the RNA control through digital PCR is underway. N1 concentrations--including those of the LOD and LOQ--reported in the current version of this work may therefore be updated in future versions to reflect the quantified concentration of the RT-qPCR standard. Nonetheless, while the approach described herein limits direct comparison of N1 concentrations to those found in other studies, it does not alter trends and comparisons across facilities examined within this study. In addition, we elected to use a pooled standard curve to quantify samples on all plates to ameliorate variability in standard preparation by different analysts from plate to plate. A description of the analysis used to motivate this decision is presented in the SI (Figure S1). The absence of contamination during RT-qPCR preparation was confirmed through no template controls included on all RT-qPCR plates. Only samples quantified above the LOQ were included in subsequent analysis. From September 8, 2020 to June 8, 2021, samples were collected from each facility on 72 sampling dates, with samples from only two dates associated with method blanks having N1 concentrations above the LOD; samples collected on these two dates were flagged as contaminated and were not included in subsequent analysis.

An attenuated bovine coronavirus (BCoV) (Calf-Guard® Bovine Rota-Coronavirus Vaccine, Zoetis) was used as a process control.^15,16^ BCoV was inoculated into samples after the pasteurization step (details provided in the SI). A one-step RT-qPCR assay, adapted from previously published assays,^15–17^ targeting the transmembrane-protein gene of BCoV was used to qualitatively assess BCoV recovery for each sample using an aliquot of the extracted RNA (primers and probes purchased from Integrated DNA Technologies). Detection of BCoV was used to confirm whether viruses were recovered in samples for which the N1 target was not detected. Additional details regarding the RT-qPCR assays, standard curves, and QA/QC procedures are provided in the SI.

### Data analysis

The concentration of the N1 RNA target in wastewater (*C*_*WW*_) was determined for each sample in units of N1 gene copies (GC)/L according to Equation 1, where *N*_*r*_ is the number of N1 GC measured by RT-qPCR, *V*_*RNA,s*_ is the volume of RNA extracted from each sample (60 µL), *V*_*RNA,r*_ is the volume of template RNA added to the RT-qPCR reaction (5 µL), and *V*_*s*_ is the volume of wastewater sample analyzed (0.04 L).

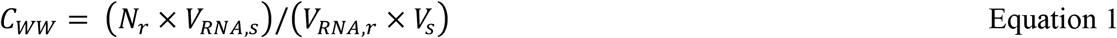

The resulting *C*_*WW*_ was then normalized by the associated daily influent wastewater flow rate (i.e., the flow rate in the same facility on the same day) to calculate the SARS-CoV-2 viral loading rate (*L*_*WW*_) in units of N1 GC/day (Equation 2). Given that 60% of the NYC sewer system is a combined stormwater-sewer system, flow-based normalization was used to account for differences in per capita water usage and variability in wastewater flow rates caused by non-domestic water inputs (e.g., rain events), which can affect measured virus concentrations. In Equation 2, *Q* is the daily flow rate at the facility in millions of gallons per day (MGD), and *CF* is the conversion factor required to convert from liters to million gallons (3.78541× 10^6^ L/MG). Continuous measurements of flow rate were conducted at each facility using either magnetic flow meters or flow measuring weirs (with uncertainty in measurements of ∼ 5%). Average daily flow rates had been measured at each facility prior to the establishment of the SARS-CoV-2 monitoring program, and thus required no additional analysis burden, making it a logistically advantageous option for normalization of virus measurements.

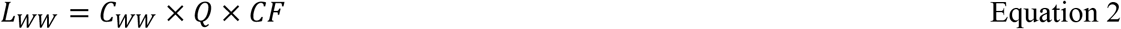

### Statistical analyses of relationships between SARS-CoV-2 loads in wastewater and laboratory-confirmed COVID-19 cases

Relationships between SARS-CoV-2 wastewater data in each sewershed and laboratory-confirmed COVID-19 cases for the associated sewershed population were evaluated through correlation and linear regression analyses. Clinical data were obtained from publicly available data provided by the NYC DOHMH.^18^ In particular, the data set “last7days-by-modzcta.csv”, which was posted online daily, was used to obtain daily reports of the cumulative clinical molecular testing results over the previous seven days for each modified ZIP code tabulation area (MODZCTA) in NYC.^18^ Specifically, data on the total clinical COVID-19 tests administered and the total number of positive tests (not including individuals who previously tested positive), reported based on date of specimen collection, were obtained. Note that molecular tests included diagnostic PCR tests and did not include antigen or antibody tests. This data set was used to calculate 7-day averages of new COVID-19 cases (i.e., positive molecular tests) per day, organized by the last date in the 7-day range. For example, the 7-day average reported on February 14 represents the daily average of new cases calculated based on the total number of positive molecular tests collected from February 8 to February 14. Data were available starting on November 7, 2020, with data from March 15, 2021 to March 21, 2021 omitted due to technical issues related to data transmission during this period (Figure S.2). While alternative data sets were available with cumulative new COVID-19 case counts prior to November 2020, these data were organized by the date that test results were reported, as opposed to date of specimen collection, and were therefore not recommended by NYC DOHMH for use in calculating the number of daily new COVID-19 cases.^18^

Each of the 177 MODZCTAs were assigned to one of NYC’s 14 sewersheds. Of the 177 MODZCTAs, 44 straddled multiple sewershed areas and were assigned to only the sewershed in which it had the greatest overlapping land area. Total new cases in each sewershed each day were calculated by summing new cases in the MODZCTA assigned to that sewershed. The same data set was used to calculate 7-day averages of COVID-19 testing rates (i.e., the number of tests administered divided by the total population) and the percentages of COVID-19 tests that were positive for each sewershed (Figure S.2).

Spearman correlations between SARS-CoV-2 viral loading rates in wastewater (N1 GC/day) and 7-day averages of new daily COVID-19 cases were determined for each individual sewershed for a five-month study period (November 8, 2020 to April 11, 2021). Correlations were also determined for a combined data set that included each data pair (i.e., SARS-CoV-2 viral loading rates and 7-day average of new COVID-19 cases on each date) for all facilities, excluding the Port Richmond and Oakwood Beach WRRFs (see the Results and Discussion section). For the combined data, correlations were also evaluated after removing data pairs associated with potentially inadequate clinical testing rates: data for dates with percentages of positive molecular tests (7-day average) that exceeded 10% in the sewershed were excluded. A general benchmark suggested by the World Health Organization in the Spring of 2020 indicated that clinical testing is less likely to represent all infections in a population when the percentage of positive tests exceeds approximately 10%;^19,20^ we therefore excluded these data in an effort to best approximate the incidence of SARS-CoV-2 infections.

To assess whether trends in SARS-CoV-2 viral loading rates in wastewater preceded trends in clinical testing data, correlations between the two data sets were also evaluated for each sewershed with the clinical data shifted back in time with lags ranging from 0 to 21 days. For this analysis, additional clinical data from April 12, 2021 to May 2, 2021 was included to maintain a constant number of data pairs for each number of lag days applied.

Simple linear regressions were performed using log_10_-transformed SARS-CoV-2 viral loading rates (N1 GC/day) and log_10_-transformed 7-day averages of new COVID-19 cases (new COVID-19 cases/day) for each individual sewershed as well as for the combined data set. The combined data set was assessed with and without the testing rate filter described above. Linear regressions were used to estimate the equivalent number of cases/day/100,000 people associated with the method LOD (*C*_*LOD*_), equal to 4,500 N1 GC/L. This estimate was calculated for each facility using individual, sewershed-specific linear regressions and using the linear regression for the combined data set. First, the LOD was converted to a SARS-CoV-2 viral loading rate in wastewater (*L*_*WW* ,*LOD*_) for each sewershed in units of N1 GC/day using Equation 3, where *Q*_*avg*_ is the average of daily flow rates at the facility over the study period (Table S.1), in MGD.

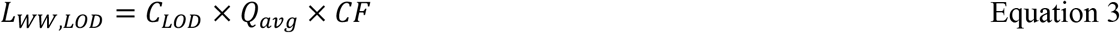

*L*_*WW* ,*LOD*_ for each sewershed were then input to the linear regressions determined for each sewershed to estimate the number of new COVID-19 cases/day associated with the SARS-CoV-2 method LOD (*Case*_*LOD*_), using Equation 4, where *m* and *b* are the slope and y-intercept of the linear regression line, respectively (presented for each sewershed in the Results and Discussion section). An example estimation is illustrated graphically in Figure S.6. Resulting *Case*_*LOD*_ values were normalized per 100,000 people using MODZCTA-level population estimates from the NYC DOHMH NYC Coronavirus Disease 2019 (COVID-19) Data.^18^

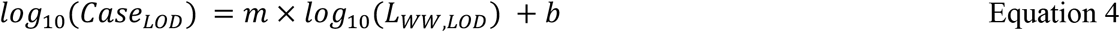

As described above, quantification of the RT-qPCR standard for the N1 target is underway. Future updates to the N1 standard concentration will change the reported method LOD, in units of N1 GC/L. However, because all sample concentrations will also be adjusted to reflect the updated standard concentration, we anticipate that the resulting relationships between the wastewater data and the clinical data (including the associated *Case*_*LOD*_) should remain similar to what is reported herein.

Statistical analyses were performed using R, and figures were created using GraphPad Prism.^21,22^

## Results and Discussion

### Methodological considerations for SARS-CoV-2 quantification in wastewater

The public health emergency caused by the emergence of COVID-19 required the expedited development of NYC DEP’s SARS-CoV-2 wastewater monitoring program. As such, several methodological choices for virus quantification were considered, and the ultimate standard operating procedure (SOP) described herein was developed reflecting NYC DEP’s program goals of monitoring trends in SARS-CoV-2 viral loads in wastewater, accounting for equipment availability, existing expertise of personnel, and considerations of material procurement. Selections were also made to minimize analyst-based variability. For example, commercially-available kits for RNA extraction were considered over alternatives that may be more sensitive to analyst skill and consistency. Data analysis and internally-developed QA/QC guidelines were established in line with programmatic goals. Additional methodological considerations, such as the inclusion of a filtration step in sample preparation, are discussed in the SI.

Long-term routine monitoring to assess virus trends through quantification with RT-qPCR requires reliable comparison of data originating from different RT-qPCR plates prepared by different analysts, which presents several challenges. First, in the absence of a formally quantified standard for the N1 RNA target, this program relied on the use of a synthetic RNA control. An approximate concentration of this RNA control was provided by the manufacturer, but was found to differ between lots purchased at different times. In addition, standard curves for routine RT-qPCR assays were prepared by different analysts on different days, with separate serial dilutions of standards performed for each individual RT-qPCR plate. To account for any resulting variability caused by these aspects of the RT-qPCR quantification method, we quantified the concentration of each RNA control lot relative to the original lot used and applied a pooled standard curve for quantification of all samples (Figure S.1). Challenges associated with RT-qPCR-based quantification using a standard curve highlight the benefits of alternative methods, such as digital PCR for absolute RNA quantification, which eliminates the need for a standard curve and may offer more sensitive detection for environmental samples.^23^ Nonetheless, the methodology employed in this work allowed us to compare relative viral loads and confidently assess of trends of SARS-CoV-2 in wastewater over time.

### SARS-CoV-2 viral loads in influent wastewater

SARS-CoV-2 viral loads in NYC’s 14 sewersheds between September 8, 2020 and June 8, 2021 were determined from quantifiable N1 gene copy (GC) concentrations in influent samples and are presented normalized by sewershed population (Table S.1^24^) in Figure 1. Maximum population-normalized SARS-CoV-2 viral loads for each facility during this period ranged from 1.6 × 10^8^ to 6.8 × 10^8^ N1 GC/day/population, with many of these values occurring around the time when a peak in COVID-19 cases was observed (January 2021). Note that in September of 2020, prior to the increase in COVID-19 cases associated with NYC’s second wave of the outbreak, N1 concentrations in wastewater remained below the LOQ in several sewersheds.

**Figure 1.**
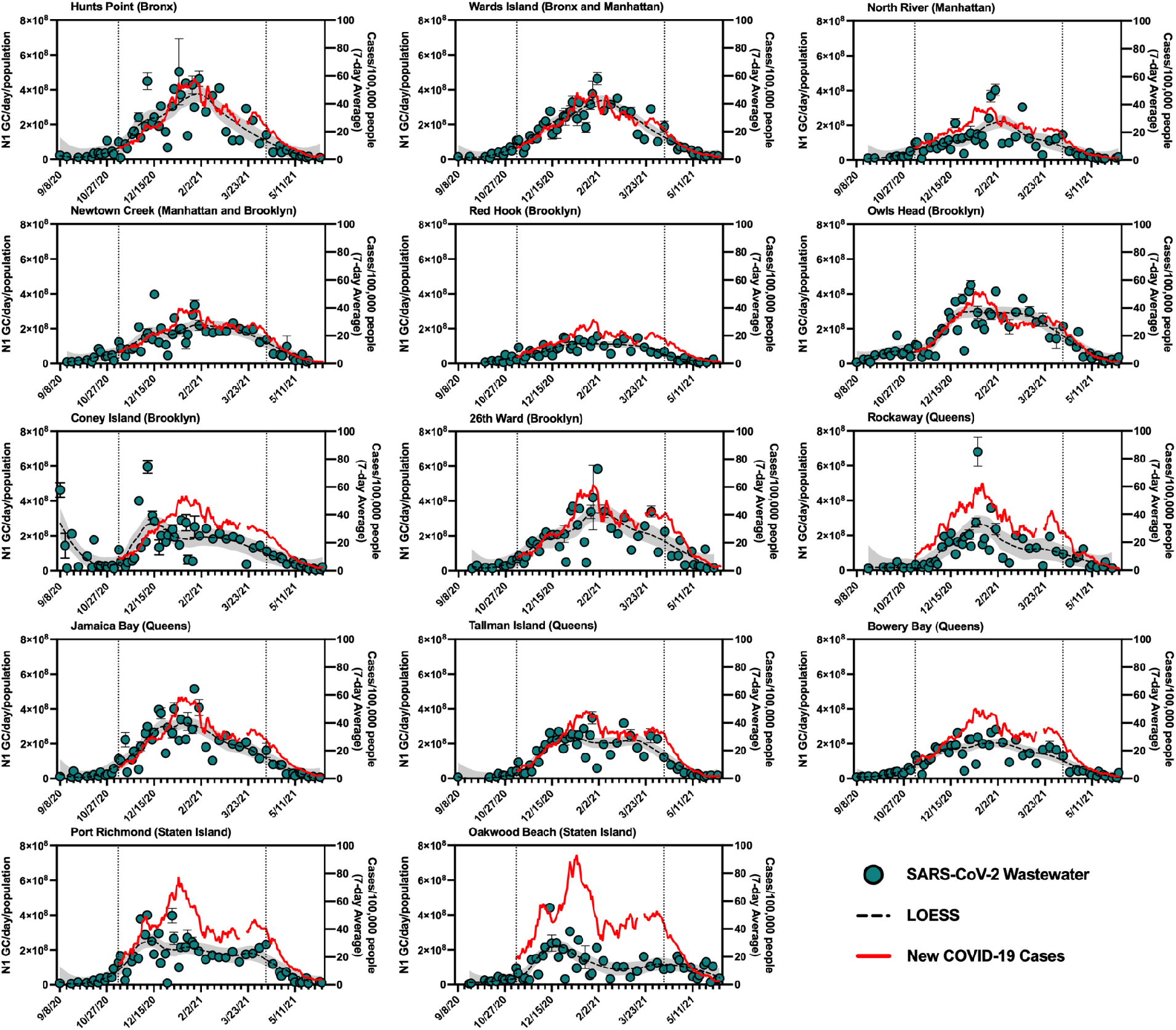
Summary of SARS-CoV-2 wastewater data for New York City’s 14 sewersheds. Data from September 8, 2020 to June 8, 2021 is shown, with the period for which statistical analysis was conducted (November 8, 2020 to April 11, 2021) bounded by vertical dotted lines. **Primary (left) y-axis, blue circles**: Influent SARS-CoV-2 viral loads normalized by sewershed populations. Error bars indicate standard deviations from triplicate RT-qPCR reactions as well as standard deviations of duplicate samples, where applicable. Dashed black lines represent LOESS curve fits (span = 0.4), with the 95% confidence intervals shaded in grey. **Secondary (right) y-axis, red line**: 7-day average of new COVID-19 cases/day/100,000 people in the previous 7 days normalized using MODZCTA-level population estimates from the NYC DOHMH’s NYC Coronavirus Disease 2019 (COVID-19) Data.^18^ Normalization by population was used for visual comparison across different sewersheds only and was not used for statistical analysis.

Visual inspection of trends in SARS-CoV-2 quantities in wastewater and new laboratory-confirmed COVID-19 cases indicates an association between the wastewater and clinical data. The strength of this association varied across sewersheds, as reflected in results from statistical analysis presented in the next section. Additionally, most sewersheds exhibited peaks for both data sets in January 2021 (Figure 1), with two notable exceptions being Oakwood Beach and Port Richmond, discussed below. Sewersheds with lower incidence rates of COVID-19 (e.g., Red Hook WRRF) generally had lower per capita SARS-CoV-2 viral loads in wastewater than those with higher incidence rates of COVID-19 (e.g., Hunts Point WRRF).

SARS-CoV-2 viral loads in the Coney Island WRRF influent in September 2020 and October 2020 displayed a high degree of variability, with some measured virus loads that were greater than those in all other sewersheds during that period, despite a consistent processing method applied for all samples and confirmed COVID-19 case rates that were consistently low across NYC (Figure 1). While there were relatively low rates of clinical testing in New York City in September 2020 and COVID-19 clusters emerged in some neighborhoods served by the Coney Island WRRF at that time,^25^ it is unclear if these factors contributed to the high viral loads measured in some Coney Island WRRF samples. For example, COVID-19 clusters were also identified in other sewersheds at this time, yet did not result in high SARS-CoV-2 loads in influent samples collected from other WRRFs, and it is difficult to determine whether clinical testing was adequate. It should also be noted that given its large geographic resolution, sewershed-level monitoring may not fully capture the effect of disease clusters (such as those identified at high spatiotemporal resolution using clinical data^26^) that may be relatively small compared to the sewershed or may straddle multiple sewersheds. Though not examined in this work, differences in wastewater quality or sewershed characteristics may also have contributed to the observed variability.

A smaller extent of variability in measured SARS-CoV-2 viral loads was observed to varying degrees across all facilities and can stem from several sources. Evaluation of duplicate samples analyzed during the study period allowed for an assessment of potential variability due to sample processing and RNA quantification. Relative standard deviations for N1 concentrations of duplicate samples (i.e., the standard deviation of concentrations from duplicate samples, each with triplicate RT-qPCR reactions, as a percent of the average concentration) ranged from 3% to 44% (mean = 17%, median = 14%); these values are comparable to those reported elsewhere for measurement of N1 concentrations in influent wastewater.^16,27^ Aside from methodological sources of variability, potential sources of variability or uncertainty include (1) dilution of wastewater from non-domestic water inputs and variations in domestic water use habits, (2) wastewater chemical composition, which may interfere with sample processing or RNA quantification methods, (3) variability in SARS-CoV-2 shedding intensity and duration for infected individuals^28–30^ and (4) the extent and consistency of viral RNA degradation in sewers.^27,31^

To account for variability in wastewater flow rates and minimize the effect of (1), viral loads calculated using measured wastewater flow rates (Equation 2) were used for analysis instead of N1 concentrations. Preliminary tests with an RT-qPCR inhibition control assay during method optimization were used to assess the impact of factor (2) and indicated minimal inhibition (data not shown). Regular assessment of inhibition with additional control assays was not feasible during routine monitoring due to resource constraints. In addition, dilution of RNA, a strategy used to reduce PCR inhibition, was avoided in order to maintain consistency in sample processing, given that viral concentrations in samples collected during periods of low COVID-19 case rates were susceptible to dilution below the limits of quantification or detection. While not included in this work, assessment of viral recovery and wastewater matrix effects should be considered for future research aiming to characterize uncertainty in WBE data. Although beyond the scope of this work, identifying and characterizing external factors related to (3) and (4) is the focus of ongoing SARS-CoV-2 WBE research efforts. Considering these uncertainties and variabilities in wastewater data, which likely increase with scale,^32^ we did not attempt to quantify the number of SARS-CoV-2 infections in each sewershed based on wastewater data, but instead explored the relationship between viral quantities in wastewater and publicly available clinical data to assess trends and associations, and examine differences between sewersheds.

As mentioned above, SARS-CoV-2 viral loads in wastewater from the Port Richmond and Oakwood Beach WRRFs (both located in the borough of Staten Island) did not capture the peak in COVID-19 cases that was observed in January 2021 across all sewersheds. In the Port Richmond and Oakwood Beach sewersheds there was a marked increase in COVID-19 cases in December 2020 that was accompanied by an associated peak in the SARS-CoV-2 viral load in wastewater during this time. However, as new COVID-19 cases in Staten Island increased by 60% in January 2021, the virus loads in wastewater stayed constant or decreased. Compared to sewersheds in the other boroughs, those in Staten Island had relatively high clinical test positivity in December and January (7-14%), despite having an average testing rate (i.e., number of clinical tests administered per capita) for the study period that was greater than that of over half of the other sewersheds (Figure S.2). This observation suggests that testing may not have adequately captured all infections in Staten Island during this period. While inadequate clinical testing rates could potentially reduce the accuracy of the observed relationships between clinical and wastewater data for these sewersheds, it does not explain the lower-than-expected SARS-CoV-2 viral loads measured in Staten Island wastewater in January 2020. A more likely explanation could stem from the composition or operation of the wastewater system in the borough. For example, a portion of the Staten Island population is not served by the sewer system and instead uses septic systems. As such, a segment of this population does not contribute to the sewer system, and viruses excreted by these residents would not have been present in the influent wastewater at the Oakwood Beach and Port Richmond WRRFs. Nonetheless, given that the population served by septic systems on Staten Island is thought to be smaller than those served by the sewer system, it is unlikely that this hypothesis can entirely explain the discrepancy between measured SARS-CoV-2 viral load and new COVID-19 cases. In addition, much of Staten Island uses separated rather than combined stormwater-sewer systems, which could potentially impact the wastewater matrix and influence viral recovery during concentration and quantification steps in sample analysis. Because of these discrepancies, the Staten Island sewersheds were excluded from analysis of the combined data set and the estimation of minimum COVID-19 case rates associated with the LOD.

By early June 2021, city-wide weekly averages of the percentage of positive COVID-19 clinical tests declined below l%, and over 50% of NYC residents had received at least one dose of a COVID-19 vaccine.^18,33^ To minimize the potential impact of mass vaccination on the evaluation of relationships between case rates and SARS-CoV-2 concentrations in wastewater presented in this work, we chose to conduct the statistical analyses described in the following section for a period ending in early April, shortly after New York State extended vaccination availability to individuals of 16 years and older.

### Relationships between SARS-CoV-2 viral loads in wastewater and new laboratory-confirmed COVID-19 cases

Significant positive correlations between SARS-CoV-2 viral loads in wastewater and new laboratory-confirmed COVID-19 cases in the corresponding populations were found for all individual sewersheds and for the combined data set (Spearman, p < 0.05), indicating, as expected, that an increase in COVID-19 cases was associated with an increase in SARS-CoV-2 concentrations in wastewater (Figure 2). Correlation coefficients (ρ) for the individual sewersheds ranged from 0.38 (Coney Island WRRF) to 0.81 (Wards Island WRRF), with an average of 0.55. Similar correlation coefficients between SARS-CoV-2 wastewater concentrations and clinical case data have been reported elsewhere.^16,34^ Note that analysis of correlations between virus concentrations (N1 GC/L, as opposed to virus loads) and new COVID-19 case rates (cases/day/100,000, as opposed to cases/day) yielded similar results (Table S.3). The correlation coefficient for the combined data set (ρ = 0.82) was higher than for any of the individual sewersheds (Figure 3.a).

**Figure 2.**
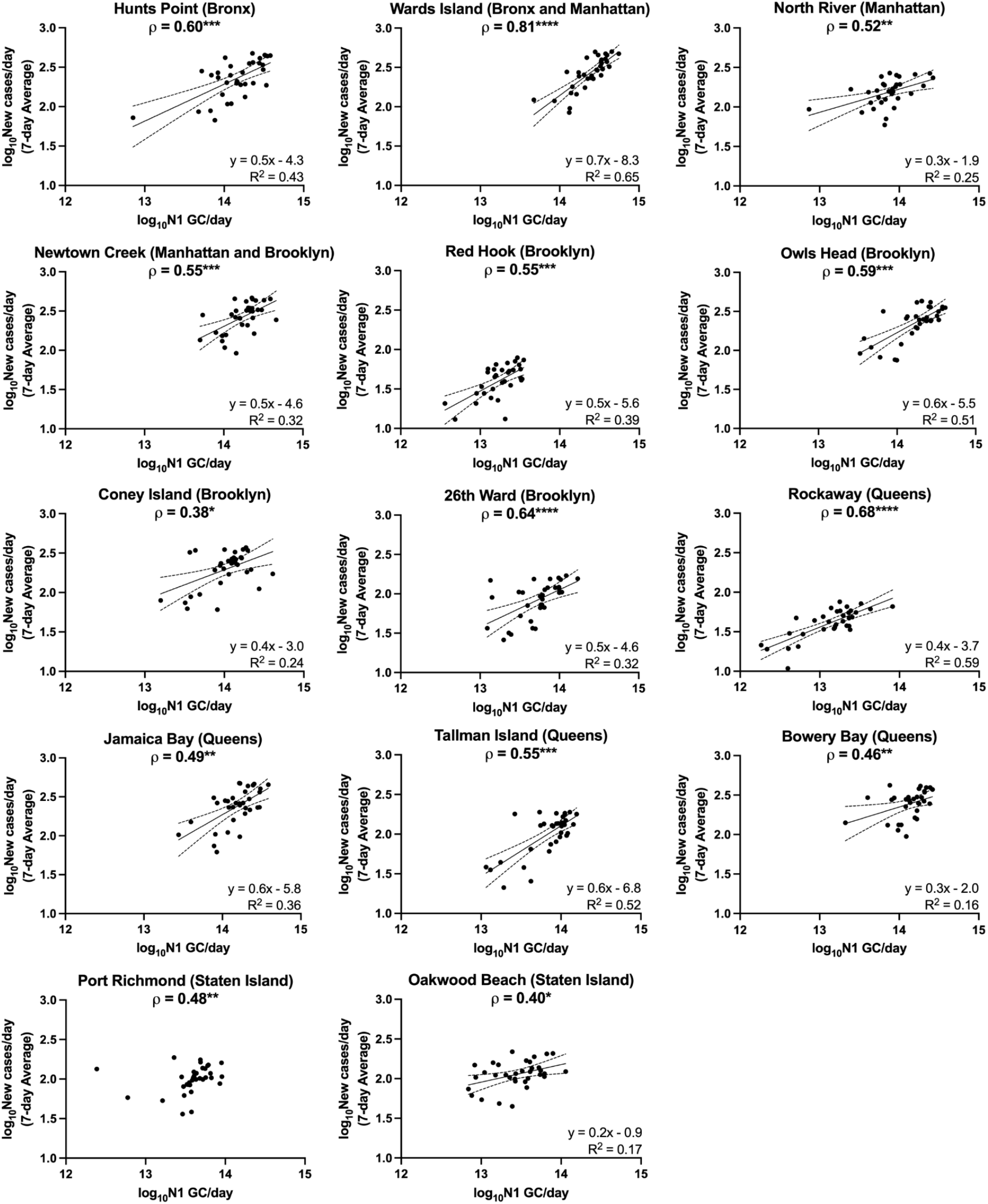
Linear regressions of log_10_-transformed SARS-CoV-2 viral loads in wastewater (N1 GC/day) and log_10_-transformed 7-day averages of new COVID-19 cases/day for each sewershed in New York City. Linear regressions (solid lines) and associated 95% confidence intervals (dashed lines) are shown along with goodness of fit R^2^ values for those data sets with significantly non-zero slopes. Note that linear regression for Port Richmond has been excluded as the slope was not significantly non-zero (see SI). The Spearman’s rank correlation coefficient (*ρ*) between N1 GC/day and new COVID-19 cases/day is shown at the top of each sewershed plot, with significance levels indicated (*p < 0.05, **p < 0.01, ***p < 0.001, ****p < 0.0001).

**Figure 3.**
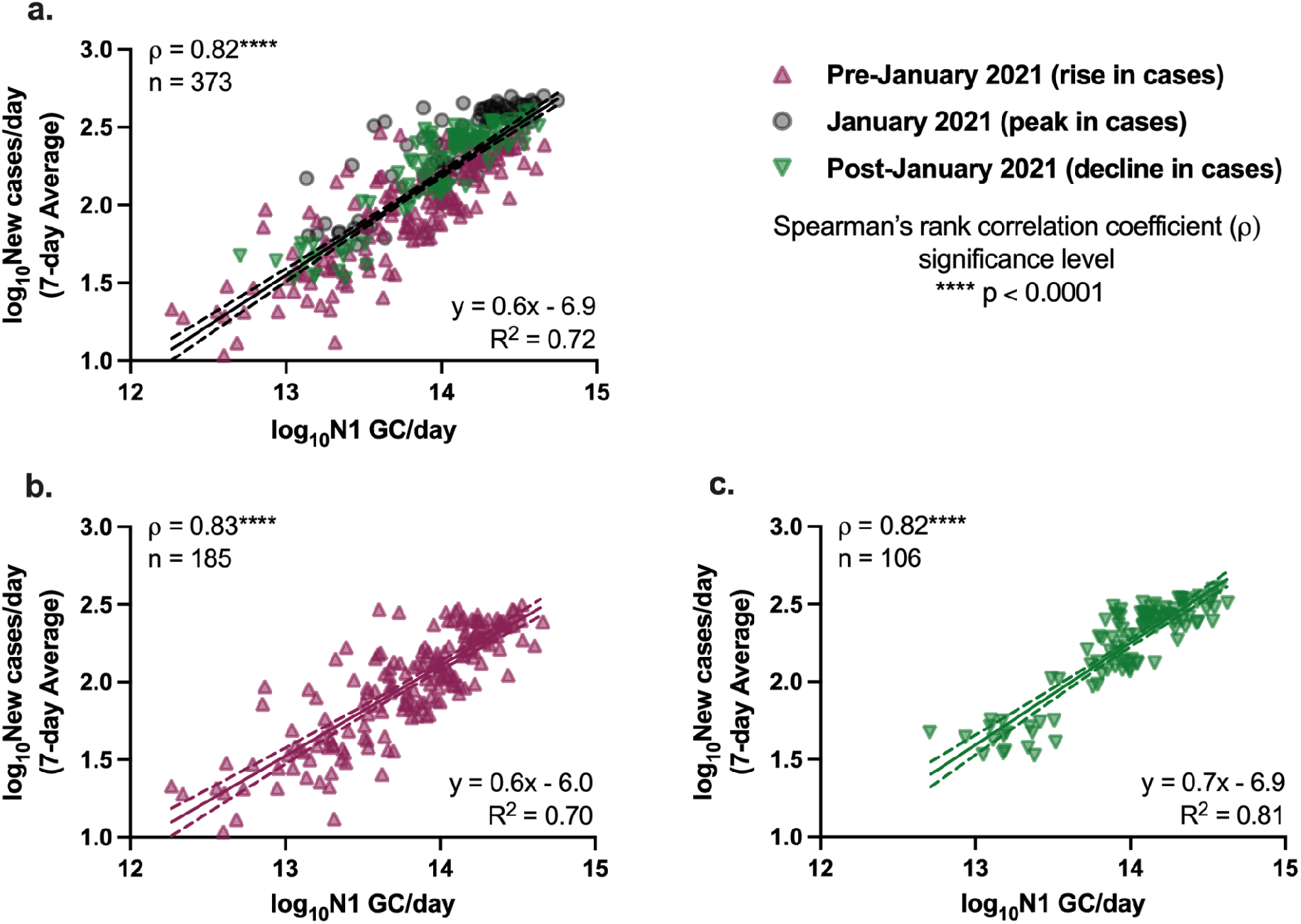
Linear regressions of log_10_-transformed flow-normalized SARS-CoV-2 viral loads in wastewater (N1 GC/day) and log_10_-transformed 7-day averages of new COVID-19 cases/day for (a) the combined data set, (b) data from the combined data set associated with a rise in cases, and (c) data from the combined data set associated with a decline in cases. Data associated with potentially inadequate testing (i.e., over 10% positive tests) are not included in this analysis. Linear regressions (solid lines) and associated 95% confidence intervals (dashed lines) are shown along with goodness of fit R^2^ values and Spearman’s rank correlation coefficients (*ρ*) between N1 GC/day and new COVID-19 cases/day.

Minimal differences were observed in the magnitudes of the Spearman’s rank correlation coefficients between clinical COVID-19 case data and SARS-CoV-2 viral loads in wastewater for the data sets with and without lag times applied (Figure S.4). Furthermore, correlations for several sewersheds--including the Wards Island WRRF--were strongest without a time lag between the two data sets. Previous studies, applying a variety of assessment methods, have suggested lag times between clinical testing and wastewater data ranging on the order of days to weeks, while others have indicated that the SARS-CoV-2 concentration in wastewater is not a leading indicator of COVID-19 diagnosis.^9^ Inconsistent findings for lag times may be attributed to whether clinical data are presented by the date of specimen collection or the date that results are reported, as well as the adequacy of COVID-19 testing rates, which vary in different regions and shift across time. Clinical data collected during periods with low testing rates are less likely to capture all infections in a region, and individuals may be more likely to be tested after symptom onset, at a time when viral shedding in feces may have already begun. These conditions can result in a lag behind wastewater monitoring data, which provides viral load information independent from clinical testing rates. Data for this work was collected during a time when testing rates were significantly higher than those during the first wave of the pandemic in NYC, and weekly median turnaround times for test results were 1 to 2 days.^18^ Furthermore, we could not confidently rule out that the small improvements in correlations observed when applying a lag time for some sewersheds was an artifact of variability in the measured wastewater data. A rigorous assessment of lag time would also need to account for contributions of previous as well as newly infected individuals to viral loads in wastewater, which was beyond the scope of this work. For these reasons, we considered data without a time lag for subsequent comparisons and linear regression analysis.

Because the nonparametric Spearman’s rank correlation was used for this analysis, results suggest that there is, at minimum, a monotonic, direct relationship between SARS-CoV-2 quantified in wastewater and clinically confirmed COVID-19 cases. Linear relationships between the two log_10_-transformed datasets were assessed through analysis of linear regressions, with the best fit found for the Wards Island WRRF (*R*^2^= 0.65) and some of the poorest fits found for the sewersheds in Staten Island (Figure 2). Inconsistent relationships between sewershed-level SARS-CoV-2 viral loads in wastewater and COVID-19 cases observed across sewersheds may be due to differences in the sewer systems for each sewershed, including sewershed areas, residence times of wastewater in the sewer system, the presence of non-domestic wastewater inputs, proportions of the population made up by transient individuals or commuters, and per capita water use. Differences could also be related to clinical testing rates for each sewershed, though no significant correlation was found between the slopes of the linear regression lines and the average testing rates for the study period for each sewershed (Spearman, p > 0.05). Similarly, no significant correlations were found between the slopes of the linear regression lines and (1) average wastewater flow rate, (2) sewershed population, or (3) average per capita wastewater flow rate (Spearman, p > 0.05), which was expected given that N1 concentrations were normalized by flow rate. Nonetheless, the linear regression found using the combined data set had a strong fit (*R*^2^= 0.70) relative to the fits of regressions for the individual sewersheds. Understanding the utility of SARS-CoV-2 wastewater monitoring data has largely involved comparison of viral concentrations in wastewater to COVID-19 case counts based on clinical testing.^35^ Because the accuracy of confirmed case rates as a measure of the number of infected individuals is dependent on COVID-19 testing rates, this comparison must be made with a consideration of clinical testing biases. Moreover, if multiple clinical data types are available, one must determine which is most appropriate for comparison to wastewater data. The analysis applied herein utilized a data set containing 7-day averages of new COVID-19 cases based on testing in each approximated sewershed area. Uncertainties surrounding such clinical testing data include (1) whether there were regional biases in testing results (Figure S.2), potentially due to testing disparities;^11^ (2) whether testing rates were adequate and what constitutes adequate testing; and (3) how long before specimen collection infected individuals contracted COVID-19 and started shedding the virus. Others have reported correlations of wastewater data with COVID-19 surveillance data sets other than clinical case rates, such as clinical test positivity or hospitalization rates.^2^ Hospital admissions data, although not without its own biases,^36^ may be an alternative epidemiological metric to compare to or to validate wastewater monitoring data if significant inadequacies in clinical testing are suspected. While hospitalization data at the MODZCTA level were not publicly available for NYC, visual comparison at the borough level indicates that trends in daily hospitalizations generally reflect trends in case rates for sewersheds within each borough (Figure S.3). The limitations of clinical testing are in fact a major driver for the application of WBE, which aims to provide community-level information free from clinical testing bias.^37–39^ Continued population-level monitoring from wastewater data could become increasingly useful in areas where clinical testing rates decline or resources for clinical testing are limited.

Linear regressions for the combined data set are presented in Figure 3 with data collected on dates with over 10% positive COVID-19 testing rates removed. Removing data associated with potentially inadequate testing from the combined data set did not significantly change the regression (Analysis of Covariance, p > 0.05) compared to the full data set without filtering (Figure S.5). After the peak in COVID-19 cases in NYC in January 2021, there was a decline in cases across all sewersheds. To assess whether the relationship between SARS-CoV-2 loads in wastewater and new clinical COVID-19 cases was significantly different during the period of declining cases from that during the period when cases were increasing, we compared separate linear regressions for the data associated with the rise in case rates (data prior to January 2021) and the decline in case rates (data after January 2021). No significant differences were found between the slopes of the linear regression lines determined using the full combined data set and the data separated based on time period.

The slope of the linear regression line for the full combined data set was found to be 0.6, indicating that a 1 log_10_ change in the number of N1 GC/day corresponded to a 0.6 log_10_ change in the number of new laboratory-confirmed COVID-19 cases/day in a sewershed. Metrics such as these are derived from relative changes in viral load, and therefore do not require absolute quantification of viral concentrations in wastewater, allowing for comparison to other studies and alleviating challenges related to absolute quantification of standard curves. However, this metric comparing SARS-CoV-2 loads and daily new COVID-19 cases has not been consistently reported in studies monitoring SARS-CoV-2 in influent wastewater. Harmonizing data analysis strategies to include such a metric would improve efforts to compare results across different locations. The slope of 0.6 observed herein is greater than that reported previously by Wolfe et al. (slope = 0.24), who compared SARS-CoV-2 concentrations measured in primary wastewater settled solids and COVID-19 incidence in seven publicly owned treatment works located across the United States, including one of the NYC facilities described in this work.^35^ In addition to analyzing a different type of sample for SARS-CoV-2 concentrations (i.e., primary settled solids versus influent wastewater), the analysis used by Wolfe et al. (2021) differed from that herein in that they normalized measured SARS-CoV-2 concentrations in wastewater solids by concentrations of pepper mild mottle virus (PMMoV). The differences in the slopes may be due to either of these factors, to variations in the relationship between SARS-CoV-2 wastewater loads and COVID-19 cases in different regions, or to a difference in the overall sensitivity of the methodology applied by Wolfe et al.

At present, limitations regarding the accuracy of COVID-19 clinical testing data and uncertainties related to SARS-CoV-2 measurements in wastewater--including SARS-CoV-2 shedding rates and RNA stability in different sewersheds--preclude development and validation of a universal, quantitative model to predict disease incidence based on viral RNA concentrations in wastewater. Ongoing research continues to expand our understanding of critical model parameters and factors contributing to uncertainty, owing particularly to SARS-CoV-2 monitoring work completed at smaller scales (e.g., building-level),^40^ from which information about the contributing population can be obtained more easily than from larger sewersheds. An attempt to quantify COVID-19 case rates in NYC’s sewersheds based on wastewater data at this time would be inaccurate, and is not currently recommended for application in the realm of public health.^41^ However, based on our analysis and others, there is utility in using wastewater data to monitor trends in COVID-19 incidence.

### Estimated case rates associated with method LOD

The utility of SARS-CoV-2 wastewater data depends on whether virions are present in wastewater at detectable concentrations (i.e., above the LOD and LOQ). It is therefore useful to approximate the minimum number of contributing COVID-19 cases per day required for detection of the SARS-CoV-2 N1 gene target in wastewater using the methodology described here. When estimated using individual, sewershed-specific linear regressions (Figure 2), the minimum new COVID-19 case rate that corresponds to the method LOD varied for each sewershed, ranging between 2 and 8 cases/day/100,000 people (Table S.4). Minimum detectable case rates were also estimated for each sewershed using the linear regression from the combined data set and the average daily influent flow rates for each WRRF during the study period. These estimates fell within the same range as those derived from sewershed-specific linear regressions (Table S.4).

The minimum detectable case rate estimates presented here should be taken as order-of-magnitude approximations rather than absolute quantities, especially considering the varying strength of the linear relationships between data for certain sewersheds (e.g., data sets for Coney Island, Bowery Bay, Oakwood Beach, and Port Richmond WRRFs had Pearson correlation coefficients below 0.5). Furthermore, these findings hold only for the specific SARS-CoV-2 quantification methodology applied herein, and may not be transferable to locations with different per capita wastewater flow rates, even if testing rates and case rates are similar to those described here. The estimates may also be limited by the assumption that the dominant source of the SARS-CoV-2 viral load in the wastewater is from recent cases as opposed to prolonged fecal shedding, which is consistent with assumptions made in previous studies.^35,42^ Furthermore, variability in virus shedding rates were not considered for the simple linear models in our study. The relationships found are also limited by the accuracy of clinical testing data, as discussed above.

As COVID-19 cases declined in NYC in the spring and early summer of 2021, the estimated minimum detectable COVID-19 case rates were reached in most sewersheds by May and June 2021. As such, we expected that SARS-CoV-2 viral loads in wastewater would have decreased to below the LOQ and LOD at this time. However, viral RNA was still detectable in influent wastewater collected from all sewersheds in mid June 2021 (Figure 4). While this discrepancy may be explained by the limitations described above, it may also be due to decreasing COVID-19 testing rates, which could result in reduced diagnosis of individuals with asymptomatic infections, who are less likely to seek out COVID-19 tests. The average COVID-19 testing rate in NYC during the period from May 2, 2021 to June 8, 2021 decreased 30% from the average in January 2021. Additionally, widespread vaccination of adults in New York may have resulted in asymptomatic and mild infections that were not diagnosed. While individuals with asymptomatic SARS-CoV-2 infections may not be captured by clinical testing, viral shedding by asymptomatic individuals would still contribute to the viral load in wastewater, given that SARS-CoV-2 has been detected in fecal samples associated with asymptomatic or mild cases of COVID-19.^43–45^ Viral loads may have also been elevated in wastewater because of prolonged fecal shedding of the virus. Finally, it is possible that the linear relationship found in this work does not hold at low SARS-CoV-2 infection levels as the study period used for statistical analysis included only case rates above the minimum detectable case rates estimated for each sewershed.

**Figure 4.**
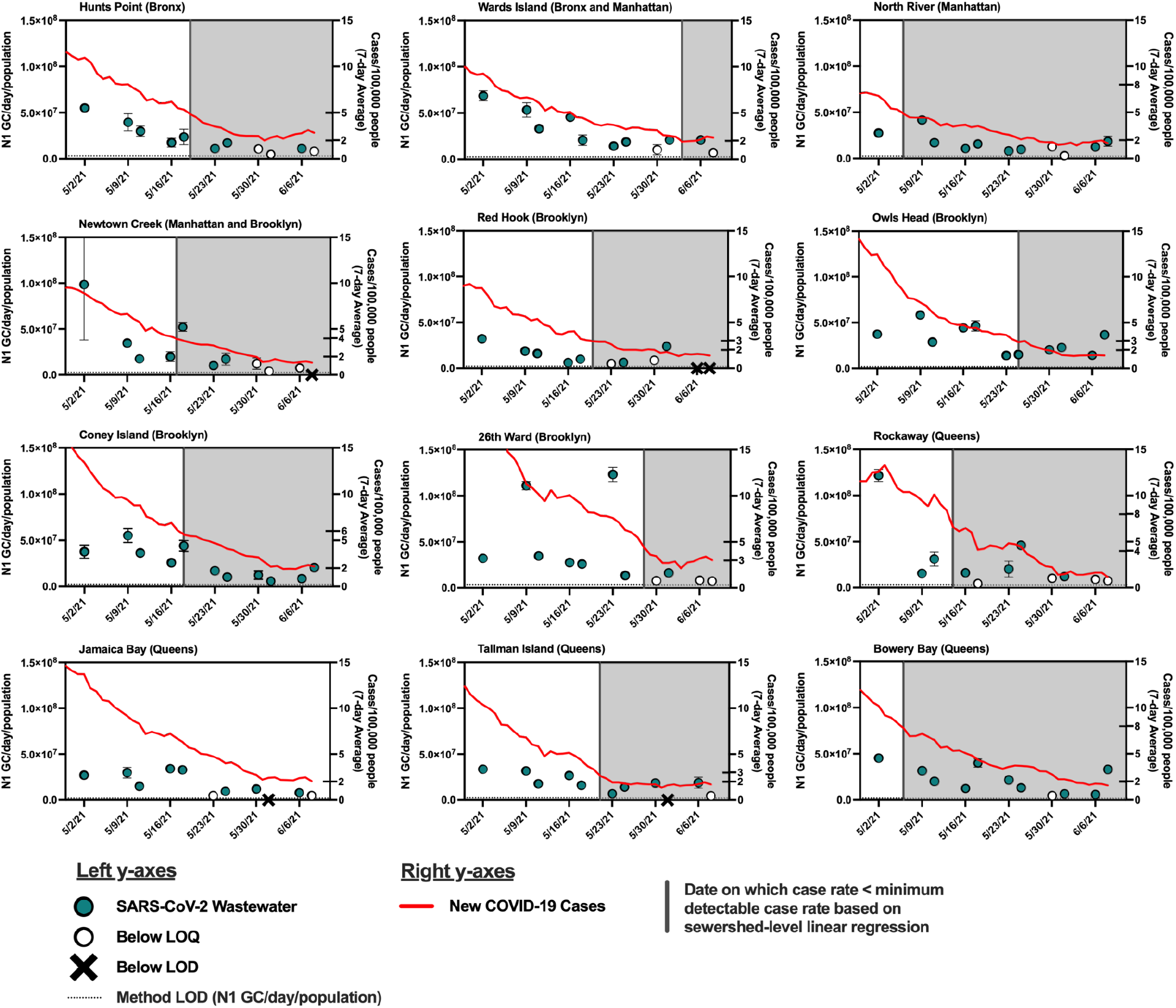
SARS-CoV-2 wastewater data and COVID-19 case data from May 2, 2021 to June 8, 2021. The date on which the case rate first fell below the estimated minimum detectable case rate (based on the sewershed-level linear regression) is indicated with a solid vertical line for each sewershed. Shaded regions indicate the time period during which case rates were below the estimated minimum detectable case rate. **Primary (left) y-axis, blue circles**: Influent SARS-CoV-2 viral loads normalized by sewershed populations. Error bars indicate standard deviations from triplicate RT-qPCR reactions as well as standard deviations of duplicate samples, where applicable. Open circles represent N1 concentrations below the limit of quantification (LOQ). Samples below the limit of detection (LOD, shown with a horizontal dotted line) are denoted with an “X.” **Secondary (right) y-axis, red line**: 7-day average of new COVID-19 cases/day/100,000 people in the previous 7 days. Estimated minimum detectable case rates (new cases/day/100,000) needed to detect SARS-CoV-2 in wastewater, based on linear regressions derived from sewershed-level data and the combined data set, are indicated with tick marks across the y-axes.

The estimated minimum numbers of COVID-19 cases required before SARS-CoV-2 can be detected in wastewater from NYC sewersheds are associated with considerable disease incidence that may be captured if some degree of clinical testing continues. Nonetheless, these estimates could aid public health agencies in understanding what COVID-19 incidence to expect if SARS-CoV-2 loads measured in wastewater influent cross the threshold from being below the detection limit to being detected. Improvements to analytical methods that lower the LOD^46–48^ would expand the utility of WBE in indicating low levels of disease incidence.

## Conclusion

Critical choices made at the beginning of the development of NYC’s SARS-CoV-2 wastewater monitoring program proved beneficial for the long-term wastewater monitoring goals for NYC, and highlight strategies that may be useful for agencies interested in implementing wastewater monitoring programs for emerging pathogens. First, collaborating parties--including academic partners and NYC DEP personnel--worked together to develop a monitoring program centered around NYC DEP’s priorities. Second, sample analysis was conducted in a NYC DEP microbiology laboratory, which allowed the program to take advantage of existing equipment, expertise, protocols, and resources related to wastewater analysis, as well as existing wastewater sampling and transport protocols and infrastructure. Doing so expedited the initiation of the wastewater monitoring program and supported virus analysis capacity building within the NYC DEP. With this structure, routine monitoring began in parallel with training and continued method optimization. Consequently, protocol adjustments responded to practical challenges as well as technical ones, taking into account laboratory infrastructure and equipment that would ultimately be used for the ongoing monitoring program. This also made for a rich training experience, in which analysts shared insights from hands-on experience, contributed to workflow decisions, and were exposed to the empirical reasoning behind methodological choices. Direct communication between wastewater treatment facility operators and laboratory personnel maximized use of the NYC DEP’s extensive knowledge base and data, which aided in troubleshooting.

As WBE programs for wastewater-related viruses evolve to meet future challenges, continued research is needed to better understand the mechanisms by which virus concentration, extraction, and quantification methods work, and the factors that influence the efficiency of each step; this knowledge can subsequently inform method optimization, standardization, and the accounting of methodological uncertainty. Since the implementation of the SARS-CoV-2 wastewater monitoring program in NYC, several studies have begun to evaluate and compare different sample processing strategies, including one interlaboratory study which included the methodology used herein.^48–50^ A clear characterization of the limitations and benefits of methodological choices for virus enumeration is critical for not only assessing previously collected data but also comparing results between WBE programs implemented by different parties, and informing future efforts in the WBE field. For example, varied priorities, resources, and expertise in different WBE programs may foster the continued use of many different methods rather than the adoption of one universal method. Additionally, poorly characterized variability in WBE data stands in the way of the critical goal of relating viral loads in wastewater to disease dynamics. Clear characterization of uncertainties related to analytical methodologies would therefore facilitate interpretation of wastewater data by public health agencies.^51^ Nonetheless, results from NYC’s monitoring program show that relative trends in SARS-CoV-2 loads in wastewater can be evaluated and associated with trends in clinical testing data, and therefore can potentially contribute to situational awareness of disease incidence in large urban sewersheds.

## Data Availability

Publicly available clinical COVID-19 datasets were retrieved from: https://github.com/nychealth/coronavirus-data. Remaining data are available upon reasonable request to the authors.

## Acknowledgements

Funding for this work was provided by the New York City Department of Environmental Protection and the Alfred P. Sloan Foundation.

An extensive team at the NYC DEP made this monitoring program possible, including Samantha MacBride, Peter Williamsen, Gina Behnke, Jasmin Torres, and Jorge Villacis; members of the NYC DEP Microbiology Lab, including William Kelly, Naudet Joasil, Patrick Hoyes, Donnovan Johnson, Manzura Kopusov, Oren Sachs, and Samantha Cruickshank; the NYC DEP transportation team, including Lateef Franklin, Samuel Young, and John Congemi; and Abeba Negatu, Patrick Jagessar, Max Verastegui and their process control laboratory teams at NYC DEP.

Several researchers at CUNY provided support and assistance for protocol development, optimization, and training, including Sherin Kannoly, Kaung Myat “Zach” San, Kristen Cheung, Anna Gao, Michelle Markman, Nanami Kubota, and Irene Hoxie.

We thank Alexandria Boehm (Stanford University) and Sandra Mclellan (University of Wisconsin-Milwaukee) for their support and guidance during program development. We also acknowledge the many insights gained from the interactions through the NSF Research Coordination Network (RCN) on Wastewater Surveillance for SARS-CoV-2.

A script automating the download of New York City’s publicly available COVID-19 clinical testing data was generously provided by Charlie Mydlarz (NYU Center for Urban Science and Progress).

## Figures

Note that the N1 concentrations reported in the following figures may be updated in future versions of this work to reflect the quantified concentration of the RT-qPCR standard, which is currently being quantified. These updates should not change observed trends reported here, as described in the main text.

