## Supplemental Information for "Monitoring SARS-CoV-2 in wastewater during New York City’s second wave of COVID-19: Sewershed-level trends and relationships to publicly available clinical testing data"

### Table of Contents

Table S.1. New York City's 14 wastewater resource recovery facilities (WRRFs) S3

**Sample Processing and RNA Extraction Methodology** S4

**RT-qPCR Assays** S6

*SARS-CoV-2 N1 Assay* S6

Table S.2. MIQE checklist: Essential and desirable information for the SARS-CoV-2 N1 target RT-qPCR assay[7] S8

Figure S.1. SARS-CoV-2 viral loads in wastewater from the Wards Island facility calculated using (a) the individual standard curves associated with the RT-qPCR plate on which each sample was run and (b) the pooled standard curve. S13

*BCoV Assay* S13

**Publicly Available Clinical COVID-19 Data and Hospitalization Data** S15

Figure S.2. Summary of COVID-19 testing data (molecular tests) for each sewershed in New York City. S16

Figure S.3. Summary of 7-day averages of new cases (solid red line) and hospitalizations (dashed black line) normalized by borough population for each New York City borough for the study period. S17

**Sewershed-level Spearman's Rank Correlation Coefficients** S18

Table S.3. Spearman's rank correlation coefficients ($\rho$) between SARS-CoV-2 wastewater data and clinical COVID-19 case data for each sewershed in New York City. S18

**Time Lag Analysis** S19

Figure S.4. Spearman rank correlation coefficients ($\rho$) between SARS-CoV-2 viral loads (N1 GC/day) and 7-day averages of new COVID-19 cases/day, with a time lag ($\tau$) between 0 and 21 days for each sewershed in New York City. S20

**Linear Regression Analysis** S21

Figure S.5. Linear regression of $\log_{10}$-transformed flow-normalized SARS-CoV-2 viral loads in wastewater (N1 GC/day) and $\log_{10}$-transformed 7-day averages of new COVID-19 cases/day for the combined data set without the data filtered based on potentially inadequate testing. S21

**Estimation of Minimum Detectable Case Rates** S22

Table S.4. Estimated minimum detectable case rates (new COVID-19 cases/day/100,000) associated with method LOD for quantification of the SARS-CoV-2 N1 gene target in wastewater (4,500 N1 GC/L) for each sewershed. S23

Figure S.6. Estimation of new COVID-19 cases/day associated with the method LOD for quantification of the SARS-CoV-2 N1 gene target in wastewater, based on the linear regression of $\log_{10}$-transformed SARS-CoV-2 viral loads (N1 GC/day) and $\log_{10}$-transformed 7-day averages of new COVID-19 cases/day for the combined data set (modified from Figure 3.a). S24

**References** S25

**Table S.1. New York City's 14 wastewater resource recovery facilities (WRRFs)**

| Wastewater Resource Recovery Facility (WRRF) | Borough(s) | Population Served* | Daily Flow Range (Average)† in MGD |
|---|---|---|---|
| Hunts Point | Bronx | 755,948 | 115 - 215 (136) |
| Wards Island | Bronx and Manhattan | 1,201,485 | 143 - 273 (180) |
| North River | Manhattan | 658,596 | 81 - 143 (94) |
| Newtown Creek | Manhattan and Brooklyn | 1,156,473 | 158 - 296 (188) |
| Red Hook | Brooklyn | 224,029 | 21 - 46 (26) |
| Owls Head | Brooklyn | 906,442 | 81 - 159 (95) |
| Coney Island | Brooklyn | 682,342 | 70 - 102 (82) |
| 26th Ward | Brooklyn | 290,608 | 44 - 89 (55) |
| Rockaway | Queens | 120,539 | 18 - 25 (20) |
| Jamaica Bay | Queens | 748,737 | 74 - 103 (81) |
| Tallman Island | Queens | 449,907 | 48 - 90 (59) |
| Bowery Bay | Queens | 924,695 | 82 - 179 (100) |
| Port Richmond | Staten Island | 226,167 | 23 - 55 (29) |
| Oakwood Beach | Staten Island | 258,731 | 24 - 41 (28) |

*Based on inter-census population estimates for 2020 from the New York Metropolitan Transportation Council's 2050 Socioeconomic and Demographic Forecast[1]
†Average ($Q_{avg}$) is based on daily flows from November 8, 2020 to April 11, 2021

**Sample Processing and RNA Extraction Methodology**

Influent wastewater samples were analyzed in 40-mL aliquots using polyethylene glycol (PEG) precipitation for virus concentration. To ensure inactivation of viruses before sample concentration, samples were first pasteurized at 60 ºC in a water bath for a total of 90 min, which allowed 30 min for the sample to reach 60 ºC and then 60 min of incubation at that temperature. Pasteurized samples were cooled in a room temperature water bath for 10 min followed by a 10 min incubation on ice prior to addition of an attenuated bovine coronavirus (BCoV) from a bovine rota-coronavirus vaccine (Calf-Guard®; Zoetis #4002), which was used as a process control. The BCoV control spike was prepared based on a method modified from Feng et al., 2021.[2] One one-dose vial of the Calf-Guard® vaccine was rehydrated with 1 mL TE buffer (Fisher Scientific, BP2473100) and stored in single-use aliquots at -80 ºC. On the day of sample analysis, an aliquot of the vaccine was thawed at room temperature and further diluted 1 in 10 using nuclease-free water. 40 µL of the diluted vaccine was added to each 40-mL sample. This spike was added after pasteurization and cooling based on preliminary analysis during protocol development that indicated reduced recovery of BCoV when it was added to the sample prior to the pasteurization step. On the contrary, we found pasteurization to increase the measured concentration of the N1 target in our samples, as compared to samples analyzed without the initial pasteurization step (data not shown).

Solids were then removed from samples through centrifugation at 5000 x *g* for 10 min at 4 ºC (Eppendorf Centrifuge 5804 R or Thermo Fisher Scientific Sorvall X4 Pro Centrifuge). Sample supernatant was filtered using 0.22 µm cellulose acetate filters (Corning, 431154). It should be noted that due to brief challenges in obtaining some consumables during the fall of 2020 due to supply chain constraints, alternative filters were used for some batches of samples—namely, (1) 0.45 µm cellulose nitrate filters (Thermo Scientific Nalgene, 130-4045PK) for samples collected on October 4, 6, and 18, 2020 and (2) 0.22 µm polyethersulfone (Millex-GP, SLGP033RS) for samples collected on November 1, 3, 8, and 10, 2020. A preliminary study indicated that filter type and size may impact virus recovery, so if filters must be used, consistency of filter type and size is preferable.

The utility of including the filtration step for sample processing and SARS-CoV-2 virus extraction depends on downstream analysis goals. For example, if extracted RNA will be used for sequencing applications, sample filtration can aid in removing bacterial cells and their nucleic acids, thereby helping to enrich for viral RNA. Preliminary work during protocol development indicated no significant difference between SARS-CoV-2 N1 gene copy concentrations in samples analyzed with and without filtration with 0.22 μm cellulose acetate filters (data not shown). Nonetheless, despite similar viral recoveries with and without filtration, we chose to include the filtration step given logistical benefits, including the prevention of clogging of membranes used in spin-column based RNA extraction methods, and ensuring that no solids were carried over after the centrifugation step. Avoiding the transfer of solids could potentially reduce variability caused by the inclusion of viruses associated with solid material, although further analysis is needed to better understand the distribution of the virus in liquid and solid fractions of wastewater samples and the impact of pasteurization on virus partitioning between these two phases.

To concentrate viruses in solution, filtered samples were added to 4.0 g of PEG (Fisher Scientific, BP233) and 0.9 g of NaCl (Fisher Scientific, BP358) in 50-mL Oak Ridge high-speed polypropylene copolymer centrifuge tubes (Thermo Scientific Nalgene, 3119-0050), shaken by hand until translucent, and held at 4 ºC overnight. Note that 50-mL Oak Ridge high-speed polycarbonate centrifuge tubes (Thermo Scientific Nalgene, 3138-0050) were initially used for sample processing; however, these tubes broke after several uses, possibly due to the polycarbonate's limited resistance to chemicals used in the RNA extraction (see below). The next day, samples were centrifuged at 12,000 x *g* for 120 min at 4 ºC (Eppendorf Centrifuge 5804 R or Thermo Fisher Scientific Sorvall X4 Pro Centrifuge) to pellet the PEG and associated virus particles.

RNA was extracted from concentrated PEG pellets using the Qiagen QIAamp Viral RNA Mini Kit (Qiagen, 52906; ethanol purchased separately, Fisher Scientific, BP2818500) following the vacuum protocol with a QIAvac 24 Plus (Qiagen, 19413), with the modifications specified here. First, 1.6x the suggested lysis buffer volume (i.e., 1.7 mL) was added directly to the PEG pellet in the Oak Ridge polycarbonate tube in which it was centrifuged to ensure recovery of the entire pellet. Note that samples processed prior to February 1, 2021 were extracted with 3x the suggested lysis buffer volume, originally used in an effort to maximize PEG pellet recovery. A subsequent study confirmed no significant difference in recovery using either 1.6x or 3x the suggested lysis buffer volume (data not shown). RNA was eluted in 60 µL of kit-supplied AVE buffer through a series of two 30 µL-elutions (Eppendorf Centrifuge 5424 R) and stored in aliquots at -80 ºC until quantification by RT-qPCR.

### RT-qPCR Assays

#### *SARS-CoV-2 N1 Assay*

A one-step RT-qPCR assay based on the CDC Diagnostic Panel was used to quantify gene copies of the N1 region of the SARS-CoV-2 (GenBank accession no. MN908947) nucleocapsid (N) gene [72-base amplicon, 28287 (starting position) - 28358 (ending position)].[3–5] Triplicate 20 µL reactions each contained 5 µL of 4x TaqPath 1-Step RT-qPCR Master Mix (Thermo Fisher Scientific, A15299); 1.5 µL of the 2019-nCoV RUO Kit primer/probe mix (Integrated DNA Technologies, 10006713) containing 6.7 µM forward primer (5′-GACCCCAAAATCAGCGAAAT-3′), 6.7 µM reverse primer (5′-TCTGGTTACTGCCAGTTGAATCTG-3′), and 1.7 µM probe (5′-FAM-ACCCCGCATTACGTTTGGTGGACC-BHQ-1-3′); 5 µL of template RNA; and 8.5 µL of nuclease-free water. Note that initial 2019-nCoV RUO Kits contained probes synthesized with Black Hole Quencher 1 (BHQ-1), while later kits contained probes synthesized with Zen/IowaBlack quenchers, according to correspondence with Integrated DNA Technologies. Thus, RT-qPCR conducted later in the study period used probes with Zen/IowaBlack quenchers. Data provided by the CDC for the limit of detection equivalence between probes with the two quencher types showed that the lowest detectable concentration at which all replicates were positive was the same for the two quencher types when using TaqPath 1-Step RT-qPCR Master Mix.[4]

Each 96-well RT-qPCR plate included triplicate no template controls (nuclease-free water). Synthetic SARS-CoV-2 RNA covering > 99.9% of the viral genome (Twist Bioscience Control 1, GENBANK ID MT007544.1; Twist Bioscience, 102019) served as both a positive control and standard used in a decimal serial dilution for quantification of N1 gene copies. Standard concentrations used for quantification ranged from 5 x $10^5$ copies/rxn (equivalent to 1.5 x $10^8$ copies/L of sample) to the limit of quantification (LOQ), which was 50 copies/rxn (equivalent to 1.5 x $10^4$ copies/L of sample). The LOQ, determined as described by Forootan et al. 2017,[6] was the concentration for which the coefficient of variation (CV) on concentrations of replicate standards calculated using measured Cq values was ≤ 35% (CV = 34% for the LOQ in this study). Note that these concentrations are relative to the approximate concentration of synthetic RNA control reported by the manufacturer, as described in the main text.

Reactions were aliquoted manually into 0.1 mL MicroAmp™ Fast Optical 96-Well Reaction Plates (Thermo Fisher Scientific, 4346907), which were covered with MicroAmp™ Optical Adhesive Films (Thermo Fisher Scientific, 4311971). RT-qPCR analysis for the SARS-CoV-2 N1 gene was conducted on a StepOnePlus Real-Time PCR System (ThermoFisher, 4376600) with the following cycling conditions: hold at 25 ºC for 2 min, 50 ºC for 15 min, and 95 ºC for 2 min, followed by 45 cycles of 95 ºC for 3 sec and 55 ºC for 30 sec. The MIQE checklist for reporting essential and desirable information[7] for the N1 assay can be found in Table S.2.

**Table S.2. MIQE checklist: Essential and desirable information for the SARS-CoV-2 N1 target RT-qPCR assay[7]**

| ITEM TO CHECK | IMPORTANCE | CHECKLIST |
|---|---|---|
| **EXPERIMENTAL DESIGN** | | |
| Definition of experimental and control groups | **E** | Provided in Materials and Methods |
| Number within each group | **E** | Provided in Materials and Methods |
| Assay carried out by core lab or investigator's lab? | D | Assay carried out by NYC DEP lab |
| Acknowledgement of authors' contributions | D | -- |
| **SAMPLE** | | |
| Description | **E** | Provided in Materials and Methods |
| Volume/mass of sample processed | D | Provided in Materials and Methods |
| Microdissection or macrodissection | **E** | N/A |
| Processing procedure | **E** | Provided in Materials and Methods |
| If frozen - how and how quickly? | **E** | N/A |
| If fixed - with what, how quickly? | **E** | N/A |
| Sample storage conditions and duration (especially for FFPE samples) | **E** | Provided in Materials and Methods |
| **NUCLEIC ACID EXTRACTION** | | |
| Procedure and/or instrumentation | **E** | Provided in Materials and Methods /SI |
| Name of kit and details of any modifications | **E** | Provided in Materials and Methods /SI |
| Source of additional reagents used | D | Provided in SI |
| Details of DNase or RNAse treatment | **E** | N/A |
| Contamination assessment (DNA or RNA) | **E** | Method blanks included |
| Nucleic acid quantification | **E** | RNA concentrations not routinely measured |
| Instrument and method | **E** | N/A |
| Purity (A260/A280) | D | N/A |
| Yield | D | N/A |
| RNA integrity method/instrument | **E** | Not determined |
| RIN/RQI or Cq of 3' and 5' transcripts | **E** | N/A |
| Electrophoresis traces | D | N/A |
| Inhibition testing (Cq dilutions, spike or other) | **E** | Discussed in Results and Discussion |
| **REVERSE TRANSCRIPTION** | | |
| Complete reaction conditions | **E** | Provided in SI |
| Amount of RNA and reaction volume | **E** | Provided in SI |
| Priming oligonucleotide (if using GSP) and concentration | **E** | Provided in SI |
| Reverse transcriptase and concentration | **E** | Provided in SI |
| Temperature and time | **E** | Provided in SI |

**Table S.2. MIQE checklist *continued***

| | | |
|---|---|---|
| Manufacturer of reagents and catalogue numbers | D | Provided in SI |
| Cqs with and without RT | D* | Not determined |
| Storage conditions of cDNA | D | N/A (one-step RT-qPCR) |
| **qPCR TARGET INFORMATION** | | |
| If multiplex, efficiency and LOD of each assay. | **E** | N/A |
| Sequence accession number | **E** | Provided in SI |
| Location of amplicon | D | Provided in SI |
| Amplicon length | **E** | Provided in SI |
| *In silico* specificity screen (BLAST, etc) | **E** | N/A |
| Pseudogenes, retropseudogenes or other homologs? | D | N/A |
| Sequence alignment | D | N/A |
| Secondary structure analysis of amplicon | D | N/A |
| Location of each primer by exon or intron (if applicable) | **E** | N/A |
| What splice variants are targeted? | **E** | N/A |
| **qPCR OLIGONUCLEOTIDES** | | |
| Primer sequences | **E** | Provided in SI |
| RTPrimerDB Identification Number | D | Not provided |
| Probe sequences | D** | Provided in SI |
| Location and identity of any modifications | **E** | N/A |
| Manufacturer of oligonucleotides | D | Provided in SI |
| Purification method | D | Not provided |
| **qPCR PROTOCOL** | | |
| Complete reaction conditions | **E** | Provided in SI |
| Reaction volume and amount of cDNA/DNA | **E** | Provided SI (one-step RT-qPCR) |
| Primer, (probe), Mg++ and dNTP concentrations | **E** | Provided in SI (TaqPath™ 1-Step RT-qPCR Master Mix, CG) |
| Polymerase identity and concentration | **E** | Provided in SI (TaqPath™ 1-Step RT-qPCR Master Mix, CG) |
| Buffer/kit identity and manufacturer | **E** | Provided in SI (TaqPath™ 1-Step RT-qPCR Master Mix, CG) |
| Exact chemical constitution of the buffer | D | Provided in SI (TaqPath™ 1-Step RT-qPCR Master Mix, CG) |
| Additives (SYBR Green I, DMSO, etc.) | **E** | N/A |
| Manufacturer of plates/tubes and catalog number | D | Provided in SI |
| Complete thermocycling parameters | **E** | Provided in SI |
| Reaction setup (manual/robotic) | D | Provided in SI |
| Manufacturer of qPCR instrument | **E** | Provided in SI |

**Table S.2. MIQE checklist *continued***

| | | |
|---|---|---|
| **qPCR VALIDATION** | | |
| Evidence of optimisation (from gradients) | D | Not determined |
| Specificity (gel, sequence, melt, or digest) | **E** | N/A |
| For SYBR Green I, Cq of the NTC | **E** | N/A |
| Standard curves with slope and y-intercept | **E** | Provided in SI |
| PCR efficiency calculated from slope | **E** | Provided in SI |
| Confidence interval for PCR efficiency or standard error | D | Provided in SI |
| r2 of standard curve | **E** | Provided in SI |
| Linear dynamic range | **E** | Provided in SI |
| Cq variation at lower limit | **E** | Provided in SI |
| Confidence intervals throughout range | D | Not provided |
| Evidence for limit of detection | **E** | Provided in Materials and Methods |
| If multiplex, efficiency and LOD of each assay. | **E** | N/A |
| **DATA ANALYSIS** | | |
| qPCR analysis program (source, version) | **E** | Provided in SI |
| Cq method determination | **E** | Provided in SI |
| Outlier identification and disposition | **E** | Provided in SI |
| Results of NTCs | **E** | Provided in Materials and Methods |
| Justification of number and choice of reference genes | **E** | N/A |
| Description of normalisation method | **E** | N/A |
| Number and concordance of biological replicates | D | N/A |
| Number and stage (RT or qPCR) of technical replicates | **E** | Provided in Materials and Methods |
| Repeatability (intra-assay variation) | E | Triplicate RT-qPCR reactions (SD included) |
| Reproducibility (inter-assay variation, %CV) | D | Not determined |
| Power analysis | D | Not determined |
| Statistical methods for result significance | **E** | N/A |
| Software (source, version) | E | Provided in SI |
| Cq or raw data submission using RDML | **D** | Not provided |

Cq values were determined based on the automatic Cq threshold assigned by the StepOne™ Software v2.3 (ThermoFisher). The mean and standard deviation of automatic Cq threshold across all plates were 0.30 and 0.065, respectively. Any RT-qPCR plates assigned an automatic Cq threshold more than two standard deviations above or below the mean automatic Cq threshold were designated as outlier Cq thresholds and reanalyzed with a manual Cq threshold set at 0.31, the mean Cq threshold calculated after the outlier Cq thresholds were removed.

An approximate concentration of the synthetic RNA control used for standard curve preparation was specified by the supplier. Concentrations of standards prepared from the first lot of this control purchased by the research team (referred to as the "original" lot) were determined assuming this approximate concentration. Data from nine standard curves generated with the original lot of synthetic RNA control (analyzed on different RT-qPCR plated on different days) were pooled to obtain one reference standard curve for the original lot. RNA target concentrations of subsequent lots of the synthetic RNA control were each different from that of the original lot, as evidenced by different Cq values of each point on the standard curve (data not shown). Because absolute quantification (e.g., using ddPCR) of the RNA control was not feasible at the time that sample analysis began, concentrations of subsequent lots of the RNA control were quantified using measured Cq values of the new lot of the RNA control and the pooled reference standard curve for the original lot (i.e., assuming the approximate quantity reported by the manufacturer). Note that quantification of the RNA control through digital PCR is underway, and N1 concentrations reported in the current version of this work may therefore be updated in future versions to reflect the quantified concentration of the RT-qPCR standard.

The quality assurance and quality control guidelines developed internally for the NYC DEP's SARS-CoV-2 monitoring program established an acceptable range of amplification efficiencies between 70% and 115%. PCR amplification efficiencies for all N1 assay plates ranged between 72% and 109%, with $R^2$ values for all standard curves ≥ 0.97. Of the 37 individual N1 assay plates from the study period (samples collected between November 8, 2020 and April 11, 2021), two resulted in efficiencies less than 85% and none resulted in an efficiency over 110%, indicating consistent acceptable performance of the assay over the five-month period of statistical analysis. It should be noted that variations in amplification efficiency--calculated based on the slope of the standard curve--may reflect errors or inconsistencies in preparation of standards rather than changes in actual PCR amplification efficiency of the assay. We considered variation in standard preparation a possibility, given that RT-qPCR plates were prepared by multiple analysts, with new serial dilutions of the standard prepared for each plate. To account for this potential variability and reduce any resulting noise in the data, we elected to apply a pooled standard curve to calculate N1 concentrations of all samples. The pooled standard curve was developed by combining data of standard curves from 56 plates (samples from September 8, 2020 to April 11, 2021); standard curves were pooled after the concentration adjustments of each lot of the RNA control described above were performed. The resulting pooled standard curve had a slope = -3.52, PCR efficiency = 92% (with 95% confidence interval of 91% to 94%), y-intercept = 41.01, and $R^2$ = 0.99. A comparison of N1 concentrations measured for the Wards Island facility using (a) individual and (b) pooled standard curves (Figure S.1) demonstrates how this approach addresses variability due to errors during standard curve preparation without affecting general trends in the data.

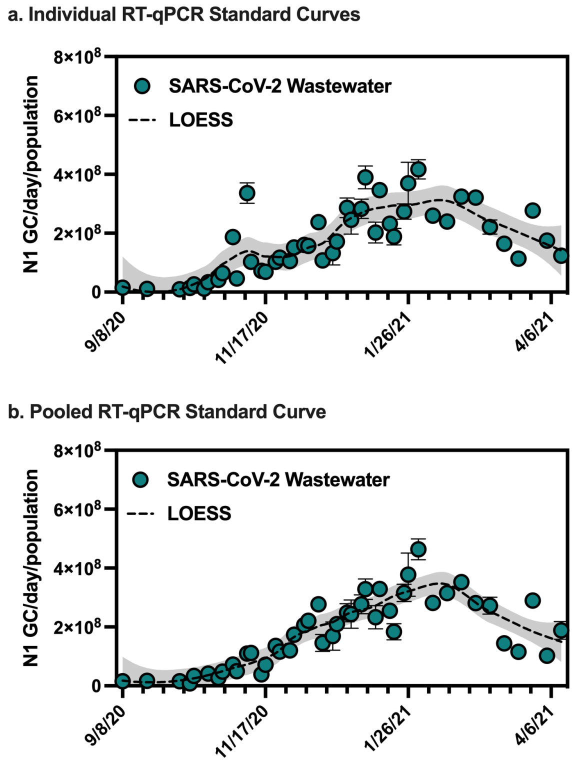


**Figure S.1. SARS-CoV-2 viral loads in wastewater from the Wards Island facility calculated using (a) the individual standard curves associated with the RT-qPCR plate on which each sample was run and (b) the pooled standard curve.**
Influent SARS-CoV-2 viral loads were normalized by the sewershed population. Error bars indicate standard deviations from triplicate RT-qPCR reactions as well as standard deviations of duplicate samples, where applicable. The dashed black line represents a LOESS fit (span = 0.4), with the 95% confidence intervals shaded in grey.

***BCoV Assay***

A one-step RT-qPCR assay adapted from previously published assays[2,8,9] targeting the transmembrane (M) gene of BCoV was used to assess recovery of the process control (primers and probes purchased from Integrated DNA Technologies). Triplicate 20 μL reactions each contained 5 μL of TaqPath 1-Step RT-qPCR Master Mix (4x, ThermoFisher), 1.5 μL of the primer/probe mix containing 5 μM forward primer (5′- CTGGAAGTTGGTGGAGTT-3′), 5 μM reverse primer (5′- ATTATCGGCCTAACATACATC-3′), and 2.5 μM probe (5′-FAM-CCTTCATAT/Zen/CTATACACATCAAGTTGTT/3IABkFQ-3′), 5 μL of template RNA, and 8.5 μL of nuclease-free water. Each PCR plate included triplicate no template controls (nuclease-free water). A custom gBlocks gene fragments oligo (Integrated DNA Technologies) (5′-

GTATCAGGTTGTTTATTAGAACTGGAAGTTGGTGGAGTTTCAACCCAGAAACAAACA ACTTGATGTGTATAGATATGAAGGGAAGGATGTATGTTAGGCCGATAATTGAGGAC TACCATACCCTTA-3′) served as both the positive control and standard used in a decimal serial dilution for quantification of gene copies. RT-qPCR analysis was conducted on a StepOnePlus Real-Time PCR System (ThermoFisher) with the following cycling conditions: hold at 25 ºC for 2 min, 50 ºC for 15 min, and 95 ºC for 12 min, followed by 45 cycles of 95 ºC for 3 sec, 55 ºC for 30 sec, and 60 ºC for 1 min.

Relative standard deviations ($RSD$) of both N1 concentrations and BCoV target concentrations for duplicate samples were calculated using equation S1, where $SD_{GC}$ is the standard deviation and $AVG_{GC}$ is the average gene copy concentration from duplicate samples, each with triplicate RT-qPCR reactions.

$$RSD = \frac{SD_{GC}}{AVG_{GC}} \times 100\% \qquad \text{Equation S1}$$

In general, the relative standard deviation for concentrations of the BCoV target were not consistent with those of the N1 target in a given sample, indicating that quantified recovery of the BCoV control inoculated into samples before virus concentrations and extraction may not accurately reflect recovery of SARS-CoV-2. Limitations of using proxy control viruses have been discussed elsewhere.[10] Calculated recoveries based on the known concentration of the BCoV control spike were therefore not used to adjust N1 gene copy concentrations. However, if the BCoV control was not recovered in any sample for which N1 was also not detected, that sample was flagged for failed processing and was excluded from trend analysis or, when possible, full analysis starting from pasteurization was repeated. If the BCoV control was detected in a sample, any non-detect wells from the N1 target assay for that sample were assigned a concentration of zero, which was used in calculating the reported average concentration from triplicate wells.

### Publicly Available Clinical COVID-19 Data and Hospitalization Data

Figure S.2. summarizes the COVID-19 clinical testing data set obtained from publicly available data provided by the NYC Department of Health and Mental Hygiene (DOHMH). Figure S.2. includes, for each sewershed, the 7-day average of (1) the percentage of positive clinical COVID-19 tests, (2) new cases/day, and (3) tests/day for the past 7 days. Note that the percentage of positive clinical COVID-19 tests calculated as described in the main text, using the "last7days-by-modzcta.csv" data set, differs from the percent positivity calculated by NYC DOHMH in publicly available data sets such as "percentpositive-by-modzcta.csv", which accounts for duplication related to an individual being tested more than once during a 7-day period.[11] The percentage of positive clinical COVID-19 tests we calculated for this analysis was used only for an estimate of adequate testing (i.e., for filtering the combined data set to remove data for dates with percentages of positive molecular tests (7-day average) that exceeded 10%) and not for direct comparison to the wastewater data. Data from March 15, 2021 - March 21, 2021 were omitted due to technical issues related to data transmission. COVID-19 case data used in correlation and linear regression analyses were not normalized by population.

Figure S.3 summarizes borough-level hospitalizations from the NYC DOHMH's publicly available "hosp-by-day.csv" file.[11] Borough populations were based on MODZCTA-level population estimates from the NYC DOHMH's NYC Coronavirus Disease 2019 (COVID-19) Data.[11] Detailed information for the publicly available datasets was retrieved from: https://github.com/nychealth/coronavirus-data.

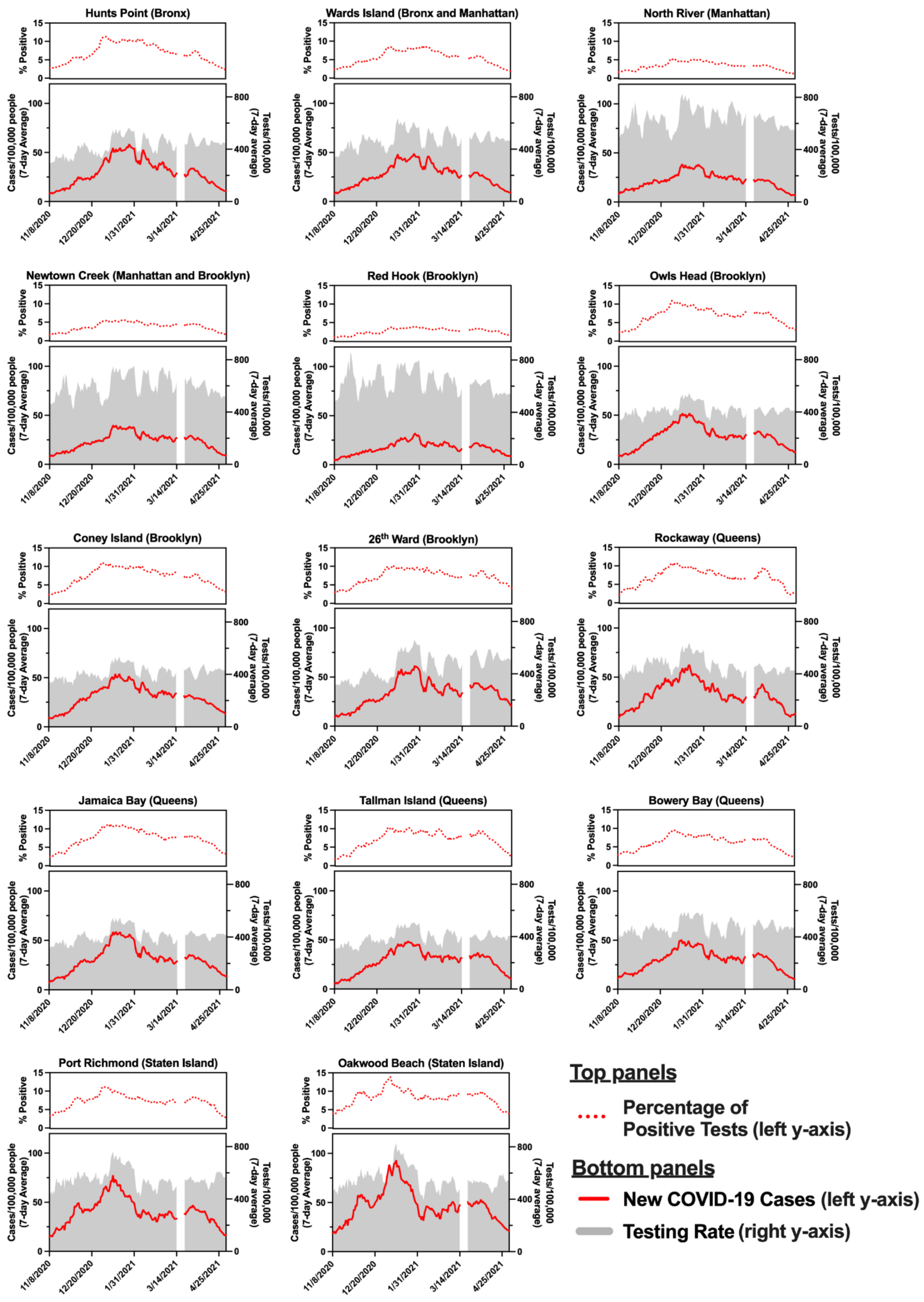


**Figure S.2. Summary of COVID-19 testing data (molecular tests) for each sewershed in New York City.**

Figure S.2. *caption continued from previous page*. For each sewershed, top panels: 7-day average of the percentage of positive clinical COVID-19 tests for the past 7 days. Bottom panels: 7-day average of new cases/day for the past 7 days (left y-axes) and 7-day average of tests/day for the past 7 days (right y-axes), both normalized by the estimated sewershed population. Note that the left and right y-axes in the bottom panels have different scales. Data used for correlation analysis described in the main manuscript text is shown (November 8, 2020 to May 2, 2021).

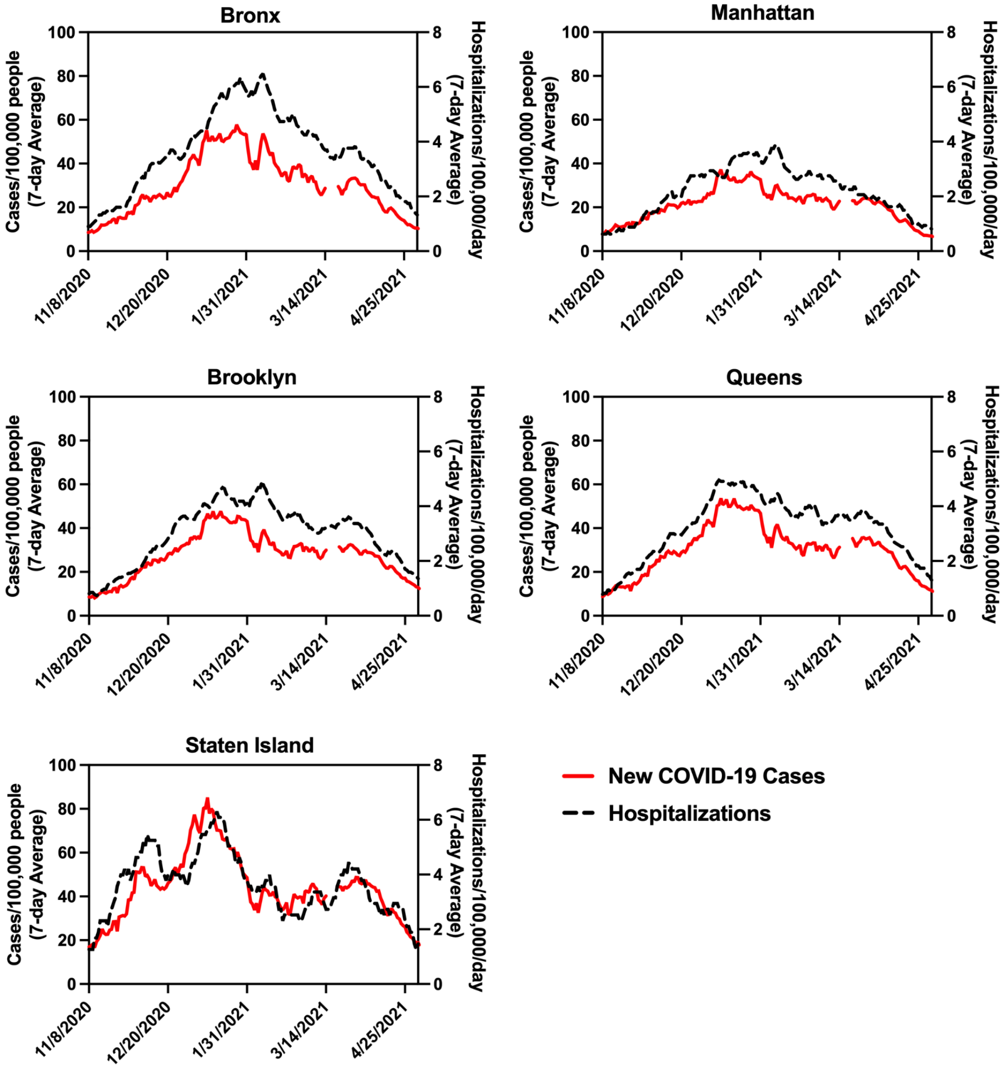


**Figure S.3. Summary of 7-day averages of new cases (solid red line) and hospitalizations (dashed black line) normalized by borough population for each New York City borough for the study period.**

Data is organized by the last date in the 7-day period for which average was calculated. Note that the left and right y-axes have different scales.

**Sewershed-level Spearman's Rank Correlation Coefficients**

**Table S.3. Spearman's rank correlation coefficients ($\rho$) between SARS-CoV-2 wastewater data and clinical COVID-19 case data for each sewershed in New York City.** Column 1: Coefficients for correlations between SARS-CoV-2 *viral loads* in wastewater (N1 GC/day) and 7-day averages of new COVID-19 cases/day, as described in the main manuscript text. Column 2: Coefficients for correlations between SARS-CoV-2 *concentrations* in wastewater (N1 GC/L) and 7-day averages of new COVID-19 cases/day normalized by sewershed populations. The alternative analysis presented in column 2 was used to assess any differences in correlation strengths due to flow normalization of wastewater data (i.e., to calculate viral loads, column 1). Significance levels: *$p < 0.05$, **$p < 0.01$, ***$p < 0.001$, ****$p < 0.0001$.

| | **Column 1** | **Column 2** |
|---|---|---|
| **Data used for correlation analysis** | **SARS-CoV-2 Viral Loads (N1 GC/day)** | **SARS-CoV-2 Concentrations (N1 GC/L)** |
| | **New COVID-19 cases/day** | **New COVID-19 cases/day/100,000** |
| Hunts Point | 0.60*** | 0.54*** |
| Wards Island | 0.81**** | 0.80**** |
| North River | 0.52** | 0.49** |
| Newtown Creek | 0.55*** | 0.52** |
| Red Hook | 0.55*** | 0.51** |
| Owls Head | 0.59*** | 0.56*** |
| Coney Island | 0.38* | 0.39* |
| 26th Ward | 0.64**** | 0.59*** |
| Rockaway | 0.68**** | 0.66**** |
| Jamaica Bay | 0.49** | 0.54*** |
| Tallman Island | 0.55*** | 0.52** |
| Bowery Bay | 0.46** | 0.38* |
| Port Richmond | 0.48** | 0.38* |
| Oakwood Beach | 0.40* | 0.41* |

**Time Lag Analysis**

To assess whether SARS-CoV-2 viral loads (N1 GC/day) measured in wastewater were a leading indicator for 7-day averages of new COVID-19 cases/day, Spearman's rank correlation coefficients between the two for lag times ranging from 0-21 days for each sewershed were assessed (Figure S.4). The time lag represents the number of days the clinical data was shifted back in time in relation to the date of wastewater sample collection. The optimal lag time (i.e., the number of days the clinical data lagged behind the wastewater data to result in the strongest correlation) varied for each sewershed, with minimal improvement in correlations associated with a lag time (Figure S.4). No significant correlations were found between the optimal lag time and the average testing rate for the study period for any sewersheds.

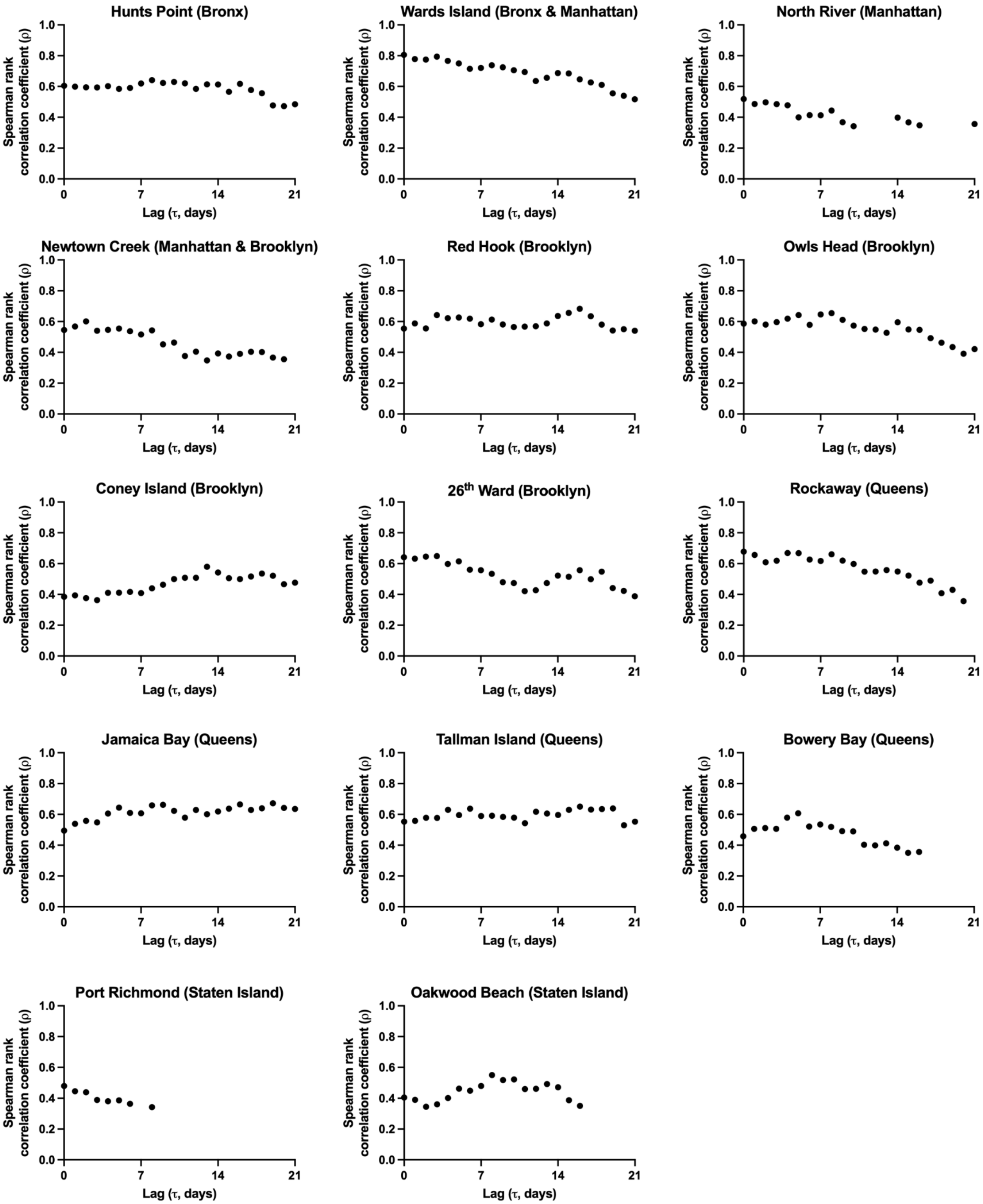


**Figure S.4. Spearman rank correlation coefficients ($\rho$) between SARS-CoV-2 viral loads (N1 GC/day) and 7-day averages of new COVID-19 cases/day, with a time lag ($\tau$) between 0 and 21 days for each sewershed in New York City.**

The time lag represents the number of days the clinical data was shifted back in time. Correlations that were not statistically significant ($p > 0.05$) have been omitted.

**Linear Regression Analysis**

Assessment of linear regressions presented in Figures 2 and 3 in the main manuscript confirmed that (1) the slope between $\log_{10}$-transformed viral loads (N1 GC/day) and $\log_{10}$-transformed new cases/day was significantly different from zero (F test; the assumption of significantly non-zero slopes held true for both the combined data set and all facilities individually, with the exception of Port Richmond), (2) a significant linear relationship was present (Pearson, $p < 0.05$; the assumption of significant linear relationships held true for both the combined data set and all facilities individually, with the exception of Port Richmond), (3) there were no clear patterns observed in residuals (though exceptions were made for some outliers which we elected to retain in the data), and (4) residuals were normally distributed based on the Shapiro-Wilks test ($\alpha$ = 0.05) considered alongside visual inspection of histograms and quantile-quantile plots. Note that linear regressions were performed using $\log_{10}$-transformed data; linear regressions with raw, untransformed data generally resulted in fits with lower $R^2$ values and more frequent cases of residuals that were not normally distributed than did regressions with the $\log_{10}$-transformed transformed data set.

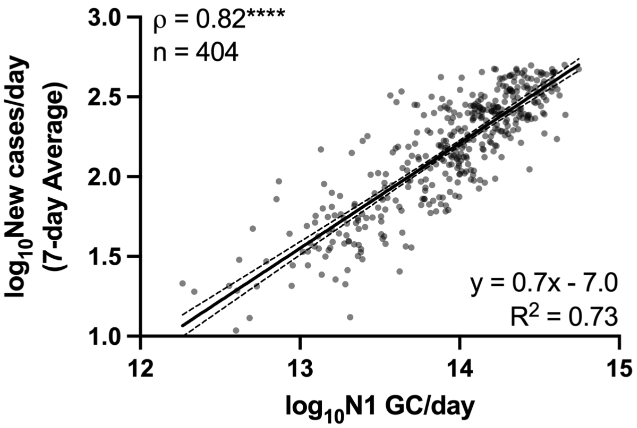


**Figure S.5. Linear regression of $\log_{10}$-transformed flow-normalized SARS-CoV-2 viral loads in wastewater (N1 GC/day) and $\log_{10}$-transformed 7-day averages of new COVID-19 cases/day for the combined data set without the data filtered based on potentially inadequate testing.**
This figure is presented for comparison to Figure 3 in the main text, which excludes data collected on dates with over 10% positive testing results. The linear regression (solid line) and associated 95% confidence intervals (dashed lines) are shown along with the goodness of fit $R^2$ value. The Spearman's rank correlation coefficient ($\rho$) between N1 GC/day and new COVID-19 cases/day is shown at the top left, with the significance level indicated (****$p < 0.0001$).

**Estimation of Minimum Detectable Case Rates**

Table S.4 summarizes the estimated minimum number of cases per day per 100,000 people in each sewershed required to detect N1 in influent wastewater based on the method LOD. Estimates were calculated using both individual linear regressions for each WRRF and the linear regression for the combined data set using Equations 3 and 4 in the main text. To assess whether the estimates calculated based on SARS-CoV-2 viral loads differed from those obtained using SARS-CoV-2 concentrations without flow normalization, estimates were also determined using linear regressions of N1 concentrations in wastewater (N1 GC/L) and new COVID-19 cases/day/100,000. The same range of estimates (2 - 8 cases/day/100,000) was obtained from linear regressions using both pairs of data sets.

**Table S.4. Estimated minimum detectable case rates (new COVID-19 cases/day/100,000) associated with method LOD for quantification of the SARS-CoV-2 N1 gene target in wastewater (4,500 N1 GC/L) for each sewershed.**

The Oakwood Beach and Port Richmond sewersheds were excluded from analysis, as described in the main text, but information for all sewersheds is included here for completeness.

| Sewershed (WRRF) | New COVID-19 cases/day/100,000 associated with method LOD | | | | | |
|---|---|---|---|---|---|---|
| | Based on linear regressions of N1 GC/day and new COVID-19 cases/day | | | | Based on linear regressions of N1 GC/L and new COVID-19 cases/day/100,000† | |
| | Sewershed-specific regressions | Regressions from combined data set* | | | Sewershed-specific regressions | Regression from combined data set |
| | | (a) all data | (b) rise | (c) decline | | |
| Hunts Point | 5 | 2 | 2 | 3 | 5 | 5 |
| Wards Island | 2 | 2 | 2 | 2 | 2 | |
| North River | 8 | 2 | 2 | 2 | 8 | |
| Newtown Creek | 4 | 2 | 2 | 2 | 5 | |
| Red Hook | 3 | 2 | 2 | 3 | 4 | |
| Owls Head | 3 | 2 | 2 | 2 | 3 | |
| Coney Island | 6 | 2 | 2 | 2 | 6 | |
| 26th Ward | 5 | 3 | 4 | 4 | 5 | |
| Rockaway | 8 | 4 | 4 | 5 | 8 | |
| Jamaica Bay | 2 | 2 | 2 | 2 | 3 | |
| Tallman Island | 2 | 3 | 3 | 3 | 3 | |
| Bowery Bay | 8 | 2 | 2 | 2 | NA‡ | |
| Port Richmond | NA‡ | 3 | 3 | 3 | NA‡ | |
| Oakwood Beach | 20 | 3 | 3 | 3 | 21 | |

*Linear regressions were determined using (a) "all data": all data from the combined data set, (b) "rise": data from the combined data set associated with the rise in case rates (data prior to January 2021), or (c) "decline": data from the combined data set associated with the decline in case rates (data after January 2021). Note that the combined data set does not include data from Port Richmond or Oakwood Beach and has been filtered to exclude data associated with over 10% positive tests.

†Estimates from these regressions are not flow-dependent; therefore, only one estimate is determined from the combined data set.

‡Slope of associated linear regression not significantly non-zero; linear regression rejected and case rate estimate not calculated

Figure S.6 illustrates graphically the approach used to estimate the equivalent number of COVID-19 cases/day/100,000 people associated with the SARS-CoV-2 quantification method LOD using linear regression for the combined data set. First, the method LOD was converted to a SARS-CoV-2 viral loading rate in wastewater (in units of N1 GC/day) for each sewershed using Equation 3 in the main text. The average daily flow rate for each WRRF ($Q_{avg}$) ranged from 21 MGD (Rockaway WRRF) to 188 MGD (Newtown Creek WRRF), resulting in a range of LOD-equivalent viral loads between 3.5 x $10^{11}$ to 3.2 x $10^{12}$ N1 GC/day across the facilities. The estimated minimum new COVID-19 cases/day required to detect SARS-CoV-2 in wastewater influent were determined by inputting this viral load into Equation 4 (main text) with the slope ($m$) and y-intercept ($b$) values from the linear regression (Figure S.6). The resulting COVID-19 cases/day (ranging from 4 to 17 new cases/day) were then normalized by the respective sewershed populations to obtain estimates ranging from 2 to 4 new COVID-19 cases/day/100,000. The same approach was applied for each sewershed-level linear regression.

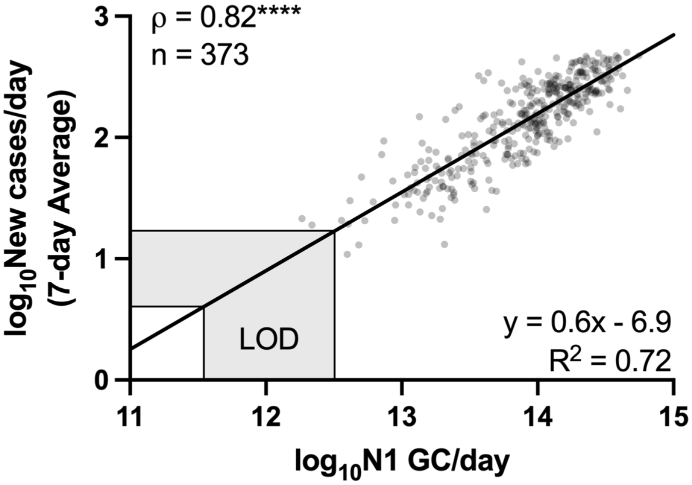


**Figure S.6. Estimation of new COVID-19 cases/day associated with the method LOD for quantification of the SARS-CoV-2 N1 gene target in wastewater, based on the linear regression of $\log_{10}$-transformed SARS-CoV-2 viral loads (N1 GC/day) and $\log_{10}$-transformed 7-day averages of new COVID-19 cases/day for the combined data set (modified from Figure 3.a).**
The method LOD (in units of N1 GC/day) for the range of average flow rates (21 MGD at Rockaway WRRF - 188 MGD at Newtown Creek WRRF) for all facilities is indicated in the shaded grey region along the x-axis. The associated minimum detectable new COVID-19 cases/day is indicated in the shaded grey region along the y-axis. Estimates from this approach were normalized by sewershed populations.
